# Compare SGLT2I versus non-SGLT2I users in type-2 diabetic mellitus patients on GLP-1 receptor agonist: A population-based and machine learning causal inference analysis

**DOI:** 10.1101/2023.11.06.23298185

**Authors:** Zhiyao Luo, Oscar Hou-In Chou, Zita Man Wai Ng, Cheuk To Skylar Chung, Jeffrey Shi Kai Chan, Raymond Ngai Chiu Chan, Lei Lu, Tingting Zhu, Bernard Man Yung Cheung, Tong Liu, Gary Tse, Jiandong Zhou

## Abstract

**Background:** Both sodium-glucose cotransporter-2 (SGLT2) inhibitors and GLP-1 receptor agonists (GLP1a) demonstrated benefits against cardiovascular diseases in type 2 diabetes (T2D). However, the effects of SGLT2I amongst patients already on GLP1a users remain unknown.

**Objective:** This real-world study compared the risks of cardiovascular diseases with and without exposure to SGLT2I amongst GLP1a users.

**Methods:** This was a retrospective population-based cohort study of patients with type-2 diabetes mellitus (T2DM) on GLP1a between 1st January 2015 and 31st December 2020 using a territory-wide registry from Hong Kong. The primary outcomes were new-onset myocardial infarction, atrial fibrillation, heart failure, and stroke/transient ischaemic attack (TIA). The secondary outcome was all-cause mortality. Propensity score matching (1:2 ratio) using the nearest neighbour search was performed. Multivariable Cox regression was used to identify significant associations. The machine learning causal inference analysis was used to estimate the treatment effect.

**Results:** This cohort included 2526 T2DM patients on GLP1a (median age: 52.5 years old [SD: 10.9]; 57.34 % males). The SGLT2I users and non-SGLT2I users consisted of 1968 patients and 558 patients, respectively. After matching, non-SGLT2I users were associated with high risks of myocardial infarction (Hazard ratio [HR]: 2.91; 95% Confidence Interval [CI]: 1.30-6.59) and heart failure (HR: 2.49; 95% CI: 1.22-5.08) compared to non-SGLT2I users after adjusting for demographics, comorbidities, medications, renal function, and glycaemic tests. However, non-SGLT2I users were not associated with the risks of atrial fibrillation (HR: 1.52; 95% CI: 0.65-3.53) and stroke/TIA (HR: 1.72; 95% CI: 0.70-4.24). The results remained consistent in the competing risk and the sensitivity analyses.

**Conclusions:** SGLT2I non-users was associated with higher risks of myocardial infarction and heart failure when compared to SGLT2I users after adjustments amongst T2DM patients on GLP1a. The result remained consistent in the machine learning causal inference analysis.

**Graphical abstract:** 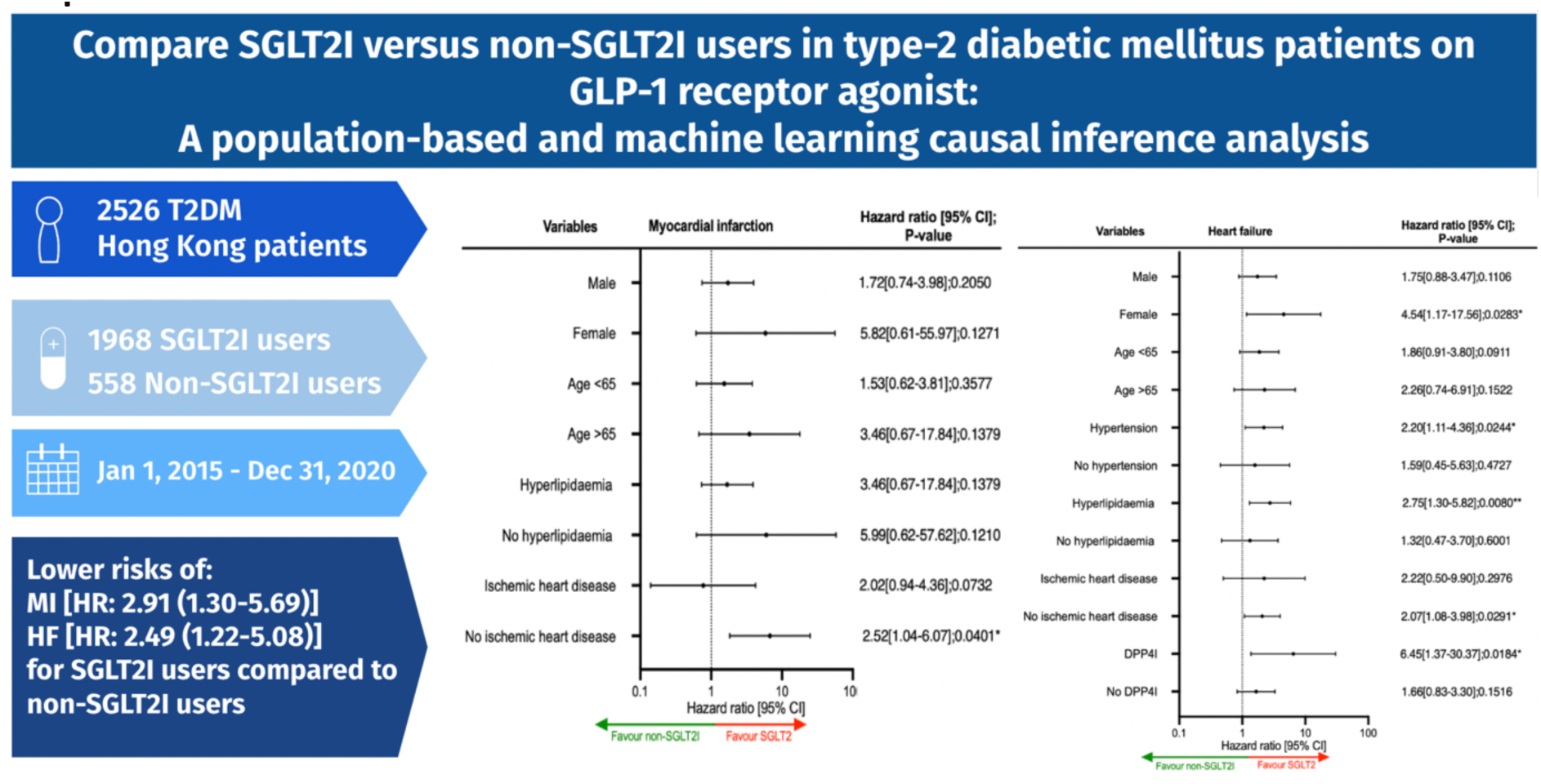

## Introduction

Cardiovascular diseases are one of the leading causes of mortality and morbidity worldwide amongst patients with type 2 diabetes mellitus (T2DM) [1]. To reduce the risks of cardiovascular diseases, current clinical guidelines highlight the importance of a multifactorial approach in diabetic management, including the use of anti-diabetic agents [2, 3]. The current literature suggests that novel anti-diabetic medications such as sodium-glucose cotransporter-2 inhibitors (SGLT2I) and glucagon-like peptide receptor agonists (GLP1a) demonstrate promising protective effects against various major adverse cardiovascular events (MACE) and mortality [4–6].

Some studies suggested that the cardiovascular benefit of SGLT2I and GLP1a is enhanced when used in conjunction [7]. A real-world cohort study demonstrates that the addition of SGLT2I amongst GLP1a diabetic users showed superior cardioprotective effects compared to the addition of sulfonylurea [8]. It was proposed that the molecular mechanism of the cardioprotective effects of SGLT2i and GLP1a could complement each other. However, the real-world evidence surrounding whether adding SGLT2I would benefit cardiovascular outcomes amongst patients using GLP1a remains limited. Previously, it was suggested that amongst patients on SGLT2I, only 4.4% of the patients had baseline GLP1a uses [9]. The recent development of machine learning (ML) for causal inference may also be applied to better characterize the treatment effects of anti-diabetic agents [10]. Therefore, this present study aims to examine the effect of SGLT2I and GLP1a on cardiovascular outcomes with ML causal inference in a T2DM cohort from Hong Kong.

## Methods

This study was approved by the Institutional Review Board of the University of Hong Kong/Hospital Authority Hong Kong West Cluster Institutional Review Board (HKU/HA HKWC IRB) (UW-20-250 and UW 23-339) and complied with the Declaration of Helsinki.

### Study population

This was a retrospective population-based study of prospectively collected electronic health records using the Clinical Data Analysis and Reporting System (CDARS) by the Hospital Authority (HA) of Hong Kong. The records cover both public hospitals and their associated outpatient clinics as well as ambulatory and day-care facilities in Hong Kong. This system has been used extensively by our teams and other research teams in Hong Kong, including diabetes and medication research.[11, 12] The system contains data on disease diagnosis, laboratory results, past comorbidities, clinical characteristics, and medication prescriptions. T2DM patients who were administered with GLP1a in centres under the HA, between 1^st^ January 2015, to 31^st^ December 2020, were included.

### Predictors and variables

Patients’ demographics include gender and age of initial drug use (baseline), clinical and biochemical data were extracted for the present study. Prior comorbidities were extracted by the *International Classification of Diseases Ninth Edition* (ICD-9) codes (**Supplementary Table 1**). The diabetes duration was calculated by examining the earliest date amongst the first date of (1) diagnosis using ICD-9; (2) HbA1c ≥6.5%; (3) Fasting glucose ≥7.0 mmol/l or Random glucose 11.1 mmol/l; (4) using anti-diabetic medications. The patients on financial aid were defined as patients on the Comprehensive Social Security Assistance (CSSA) scheme, higher disability allowance, normal disability allowance, waiver, and other financial aid in Hong Kong. The number of hospitalisations in the year prior to the index days was extracted. The Charlson standard comorbidity index was calculated. [13] The baseline laboratory examinations, including the glucose profiles and renal function tests, were extracted. The estimated glomerular filtration rate (eGFR) was calculated using the abbreviated modification of diet in renal disease (MDRD) formula. [14] Furthermore, the time-weighted lipid and glucose profiles after drug initiation were also calculated by the products of the sums of two consecutive measurements and the time interval, then divided by the total time interval, as suggested previously.[15]

### Study outcomes

The primary outcome of this study was new-onset PAD defined using ICD-9 code enlisted in **Supplementary Table 1**. Mortality data were obtained from the Hong Kong Death Registry, a population-based official government registry with the registered death records of all Hong Kong citizens linked to CDARS. Mortality was recorded using the *International Classification of Diseases Tenth Edition* (ICD-10) coding. The as-treat approach was adopted which patients were censored at treatment discontinuation or switching of the comparison medications. The endpoint date of interest for eligible patients was the event presentation date. The endpoint for those without primary outcomes was the mortality date or the end of the study period (31^st^ December 2020).

### Statistical analysis

Descriptive statistics are used to summarize baseline clinical and biochemical characteristics of patients with and without SGLT2I use. For baseline clinical characteristics, continuous variables were presented as mean (95% confidence interval/standard deviation), and the categorical variables were presented as total numbers (percentage). Propensity score matching generated by logistic regression with a 1:2 ratio for SGLT2I users versus non-users based on demographics, prior anti-diabetic drugs, number of prior anti-diabetic drugs, prior comorbidities, renal function, duration from T2DM diagnosis initial drug exposure, HbA1c and fasting glucose were performed using the nearest neighbour search strategy with a calliper of 0.1 (**Figure 1**). Propensity score matching was performed using Stata software (Version 16.0). Baseline characteristics between patients with and without SGLT2I use before and after matching were compared with absolute standardized mean difference (SMD), with SMD<0.20 regarded as well-balanced between the two groups.

**Figure 1.**
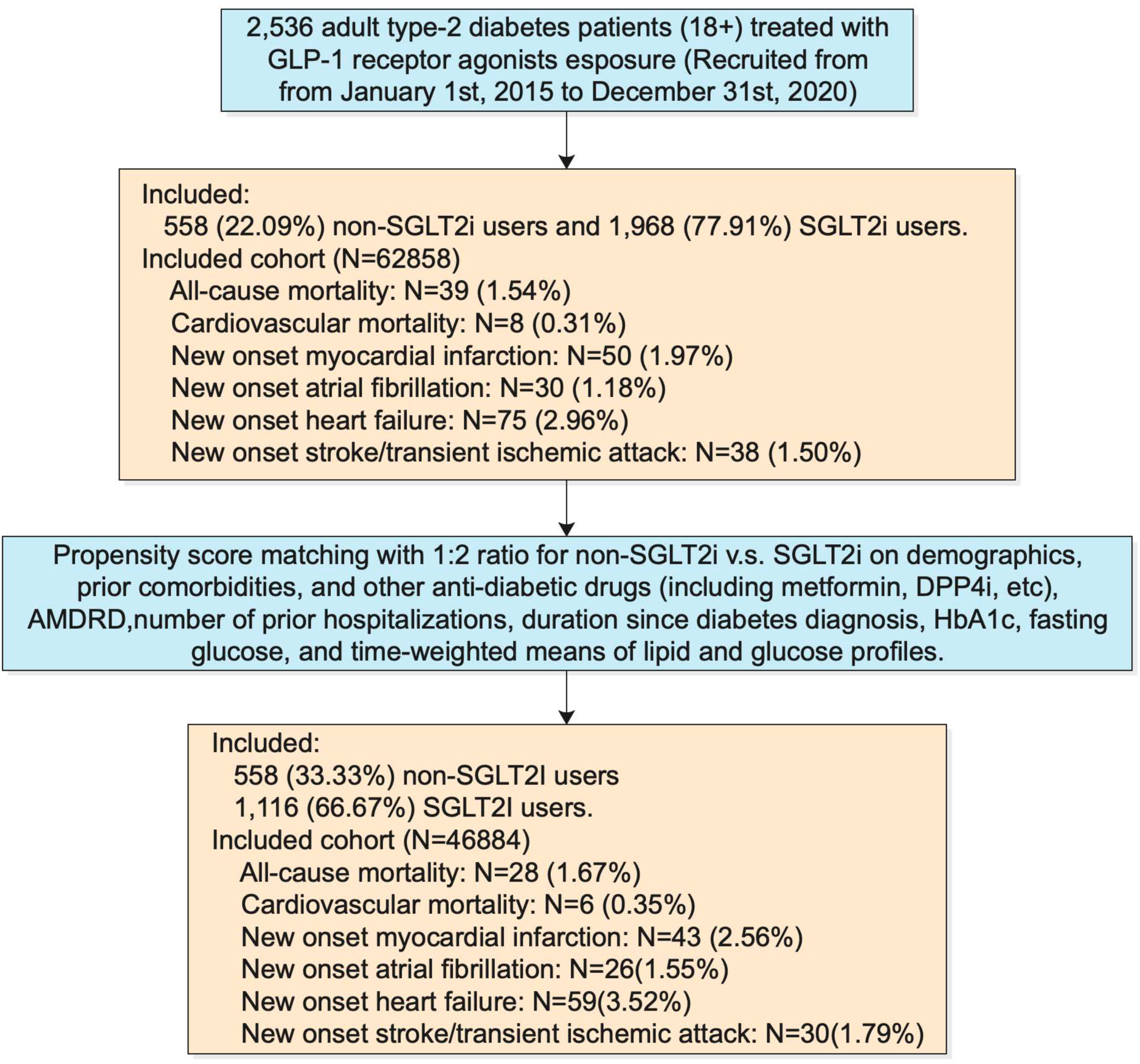
Procedures of data processing for the study cohort. SGLT2I: Sodium-glucose cotransporter-2 inhibitors; MDRD: modification of diet in renal disease.

The cumulative incidence curves for the primary outcomes and secondary outcomes were constructed. Cox proportional hazards regression was used to identify significant risk predictors of adverse study outcomes in the matched cohort, with adjustments for demographics, comorbidities, number of prior hospitalisations, medication profile, renal function, glycaemic tests, and the duration of T2DM. The log-log plot was used to verify the proportionality assumption for the proportional Cox regression models. Subgroup analyses were conducted to confirm the association amongst patients with different clinically important predictors accounting to the diabetic and the metabolic profile, as well as the comorbidities and medications associated with the outcome.

Cause-specific and sub-distribution hazard models were conducted to consider possible competing risks. Multiple propensity adjustment approaches were used, including propensity score stratification,[16] propensity score with inverse probability of treatment weighting (IPTW) [17] and propensity score with stable inverse probability weighting.[18]. In the sensitivity analysis, patients with less than 3-month follow-up duration, patients with chronic kidney disease (CKD) stage 4/5 (eGFR <30 mL/min/1.73m^2), patients on financial aid, and patients at the top or bottom 5% of propensity score matching were excluded to test the robustness of the association. We used the hip fracture as the negative control in the falsification analysis, such that the observed significant association in the falsification analysis should be attributed to bias. The hazard ratio (HR), 95% CI, and P-value were reported. Statistical significance was defined as P-value <0.05. All statistical analyses were performed with RStudio (Version: 1.1.456) and Python (Version: 3.6).

### Causal Random Forest Model

In clinical research, discerning causal links between treatments and outcomes is essential. While prediction models offer insights into probable outcomes based on various factors, they do not capture intervention causal effects[19], which is crucial distinction when assessing intervention efficacy [20].

Causal Random Forests (CRFs) stand at the confluence of machine learning and causal inference, leveraging the robust capabilities of random forests to specifically estimate heterogeneous treatment effects [21]. CRFs adeptly uncover intricate, non-linear dynamics between treatments and outcomes. CRFs manage high-order interactions, pinpointing patient subgroups deriving maximum or minimum benefit from a treatment.

Let *Y*(*t*) be the potential outcome at time *t* under treatment *A*, where *A* can be 0 (no SGLTI-2) or 1 (SGLTI-2). Our objective is to estimate the Conditional Average Treatment Effect (CATE) for time *t*:

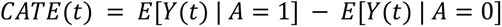

For a given patient *i*, let *W_i_* represent its covariates, *A_i_* its treatment status, and *Y_i_*(*t*) the observed outcome at time *t*. In CRFs, each tree *b* in the forest aims to predict the potential outcome differences *Y_i_*(*t*) for treatment versus control. The splitting criterion in CRFs is based on maximizing the difference in treatment effects between the two child nodes. Specifically, the impurity decrease in a node split is calculated as:

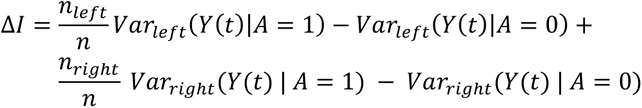

where *n* is the total number of observations in the current node, *n_left_* and *n_right_* are the number of observations in the left and right child nodes, and *Var_left_* and *Var_right_* are the variances of the potential outcomes in the left and right child nodes, respectively.

In essence, the CRF optimization process is driven by finding splits that result in nodes with high differences in treatment effects, which, in turn, reveals subgroups with varying responses to treatment. Once the forest is constructed, the CATE for a new observation *x* at time *t* is estimated as:

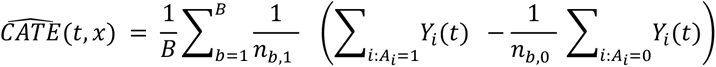

where *B* is the number of trees in the forest, *n_b_*_,1_ and *n_b_*_,0_ are the number of treated and control observations, respectively, in tree *b* for which *x* falls in the terminal node.

## Results

In this territory-wide cohort study of 2526 patients (median age: 52.5 years old [SD: 10.9]; 57.34 % males) with T2DM treated with GLP1a between 1^st^ January 2015 and 31^st^ December 2020 in Hong Kong, patients were followed up until 31^st^ December 2020 or until their deaths (**Figure 1**). This study included 1068 patients (77.91%) who used SGLT2Is, and 558 patients (22.09%) who did not use SGLT2I (Table 1). Before matching, fewer SGLT2 users had hypertension and were on less medications (metformin, anticoagulants, ACEI/ARB, diuretics).

**Table 1.**
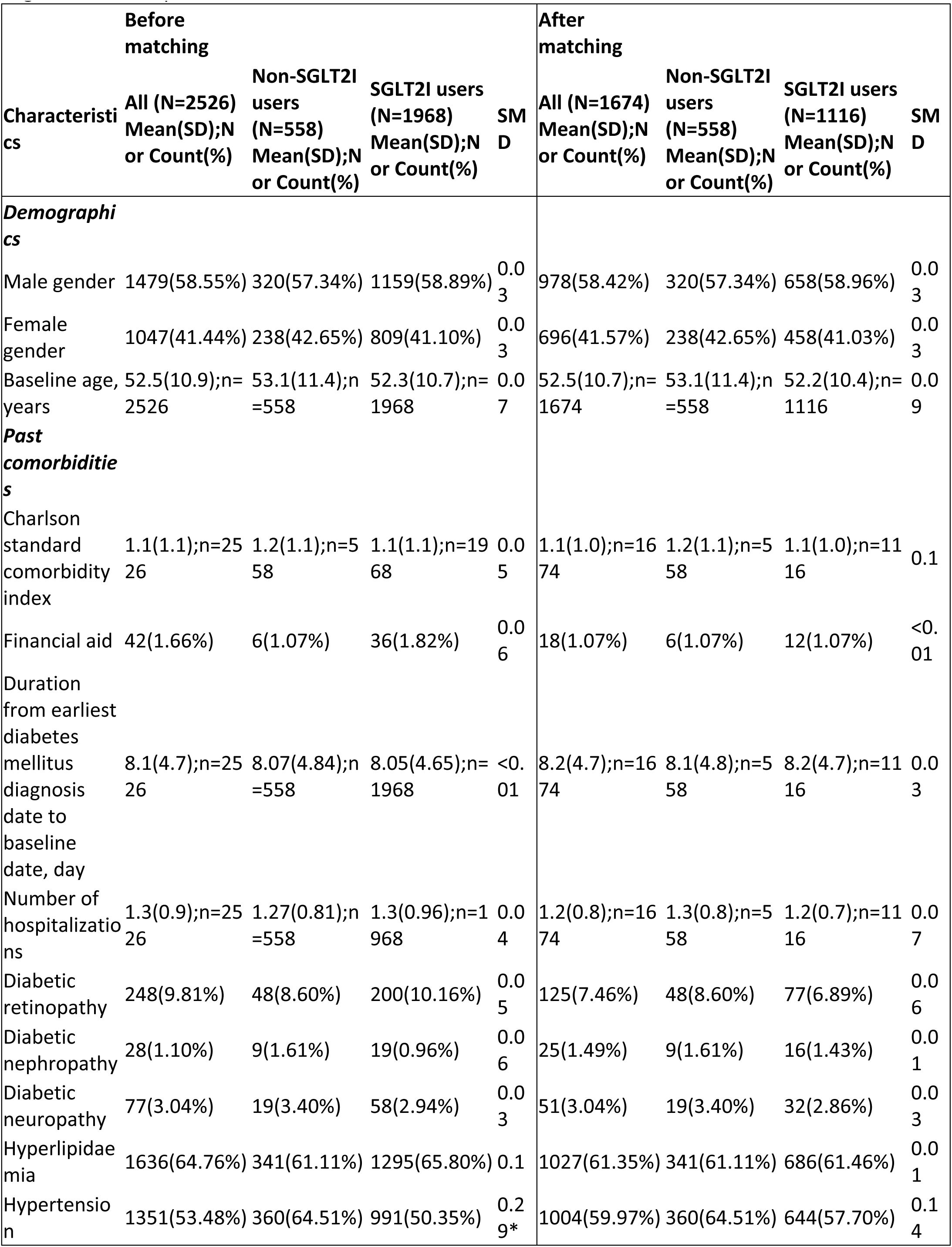
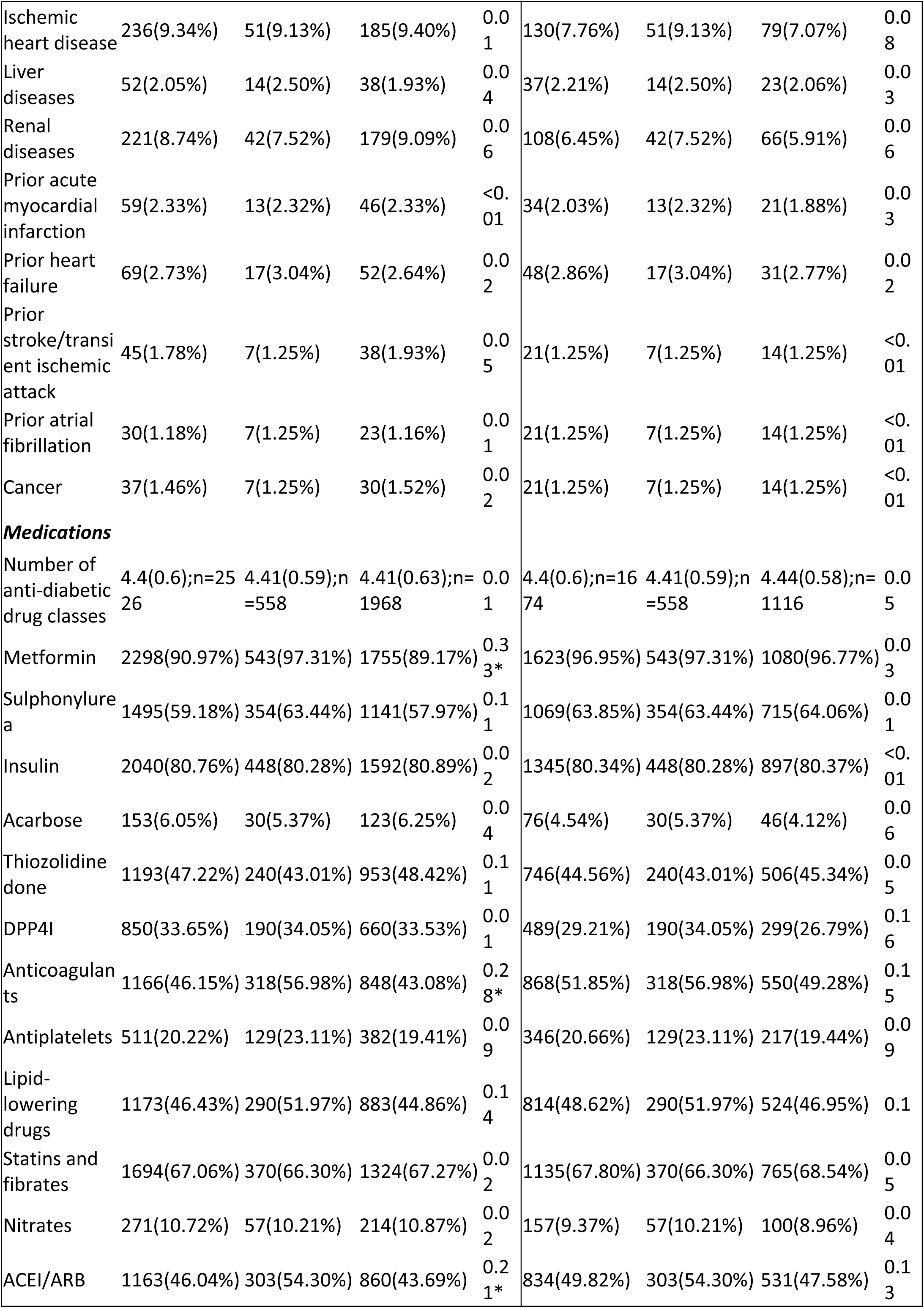
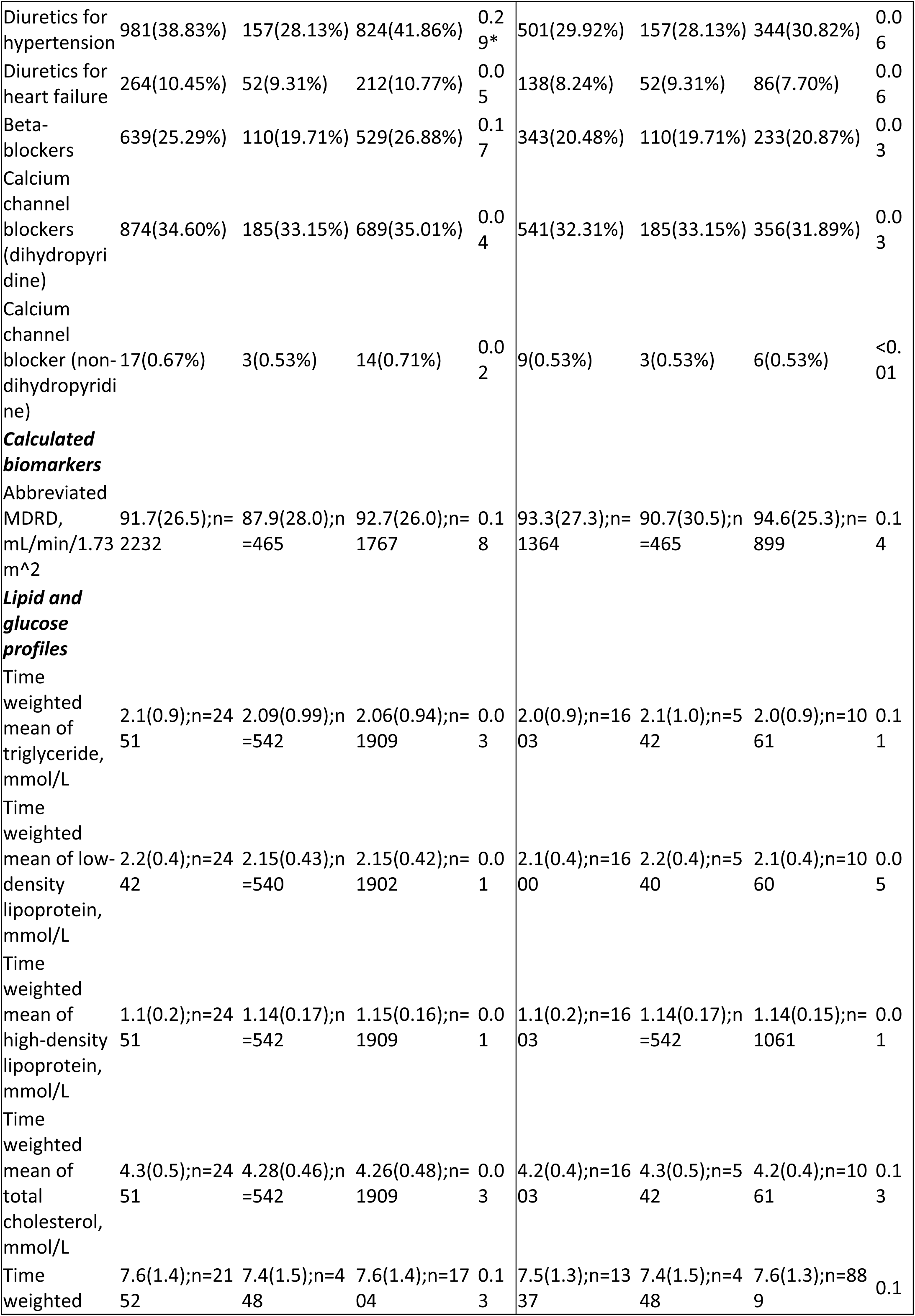
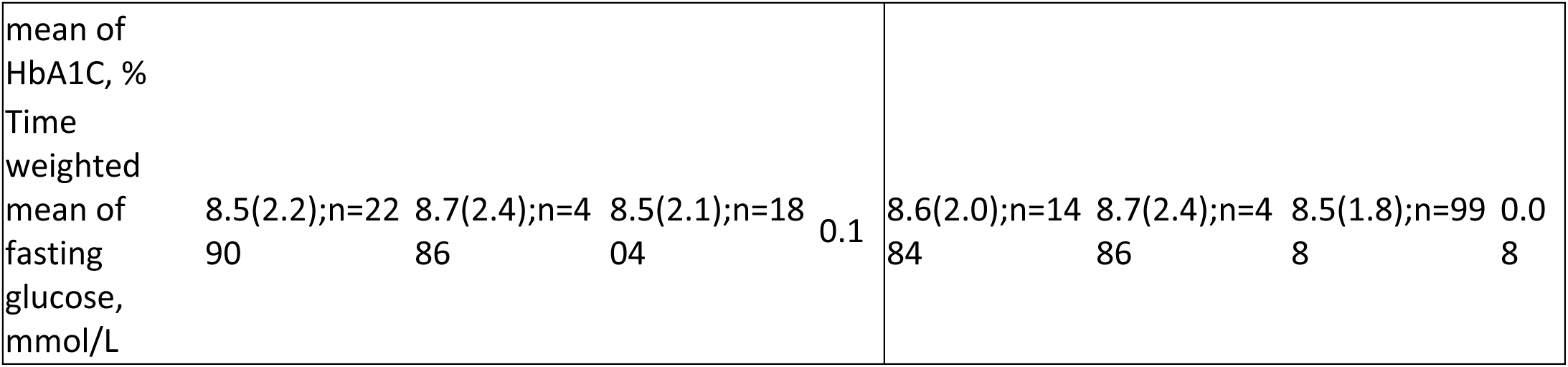
Baseline and clinical characteristics of patients with SGLT2I users versus SGLT2I non-users amongst GLP1A users before and after propensity score matching (1:2). * for SMD≥0.1; SGLT2I: sodium glucose cotransporter-2 inhibitor; DPP4I: dipeptidyl peptidase-4 inhibitor; MDRD: modification of diet in renal disease; ACEI: angiotensin-converting enzyme inhibitors; ARB: angiotensin II receptor blockers.

After the propensity score matching, the baseline characteristics and the time-weighted lipid and glucose profiles of the two groups were well-balanced **(Table 1)**. The SGLT2I and non-SGLT2I users were comparable after matching with the nearest neighbour search strategy with a calliper of 0.1, and there was no violation of the proportional hazard assumption **(Supplementary** Figure 1). In the matched cohort, 43 patients developed myocardial infarction (2.56%), 26 patients developed atrial fibrillation (1,55%), 59 patients developed heart failure (3.52%), and 30 patients developed stroke/TIA (1.79%). The characteristics of patients by drug use before and after propensity score matching are shown in **Table 1**.

### Association between SGLT2I and non-SGLT2I users and cardiovascular outcomes

In the matched cohort, 21 SGLT2I users and 22 non-SGLT2I users developed myocardial infarction. After a total follow-up of 9176.7 person-year, the incidence of myocardial infarction was lower amongst SGLT2I users (Incidence rate [IR] per 1000 person-year: 3.4; 95% CI: 2.1-5.2) compared to non-SGLT2I users (IR per 1000 person-year: 7.3; 95% CI: 4.6-11.0) **(Table 2)**. Non-SGLT2I users had a higher risk of myocardial infarction after adjustment (HR: 2.91; 95% CI: 1.30-6.50, p=0.0092) compared to SGLT2I users regardless of the demographics, comorbidities, medication profile, renal function, glycaemic tests, number of hospitalisations, and the duration of T2DM **(Table 2)**. This was substantiated by the cumulative incidence curves (**Figure 2**). The E-value for the myocardial infarction was 5.27, which suggested that the unobserved confounding variable with at least a 5.26-fold stronger association with myocardial infarction would be needed to explain the significant HR, but weaker confounding could not do so (**Supplementary** Figure 4).

**Figure 2.**
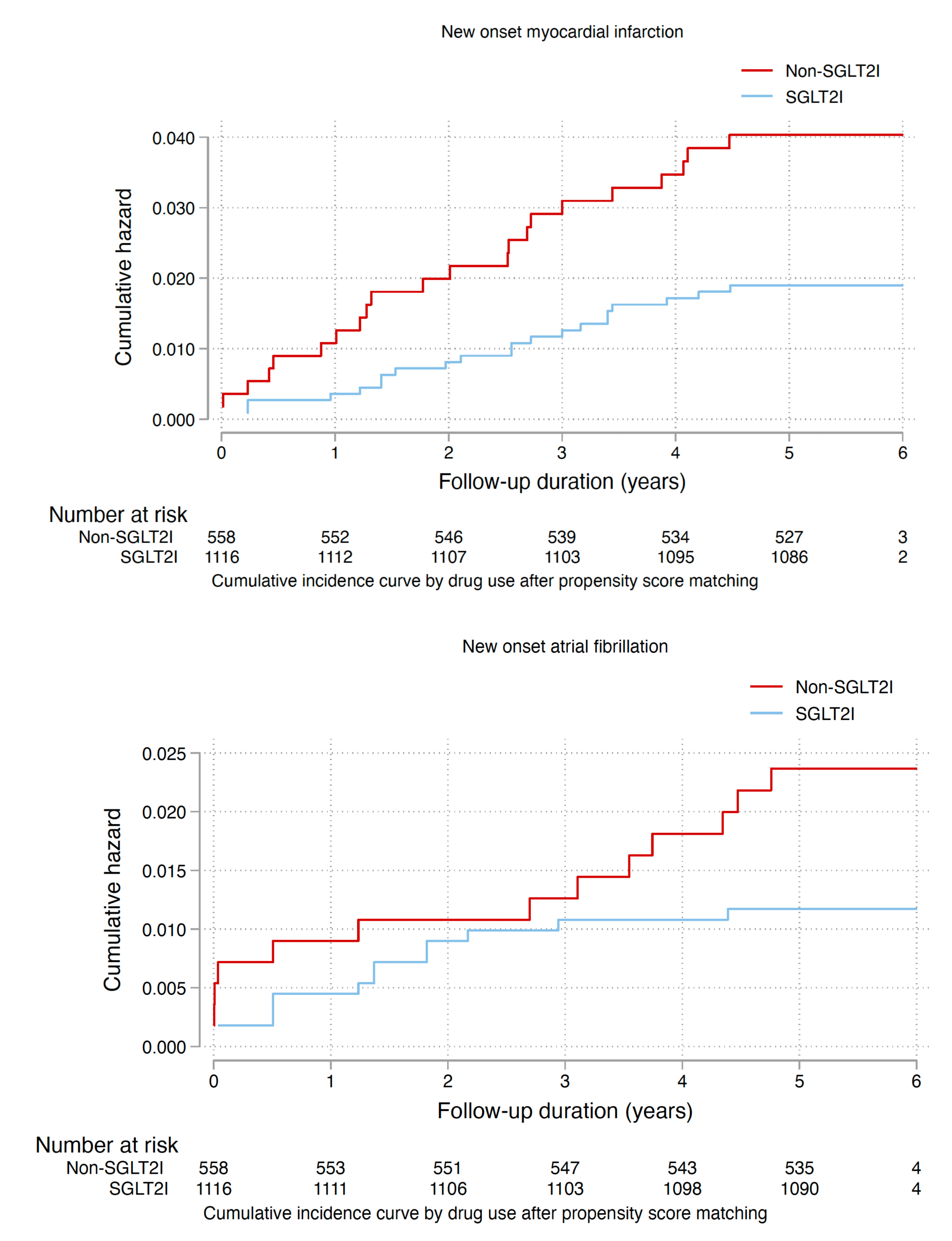
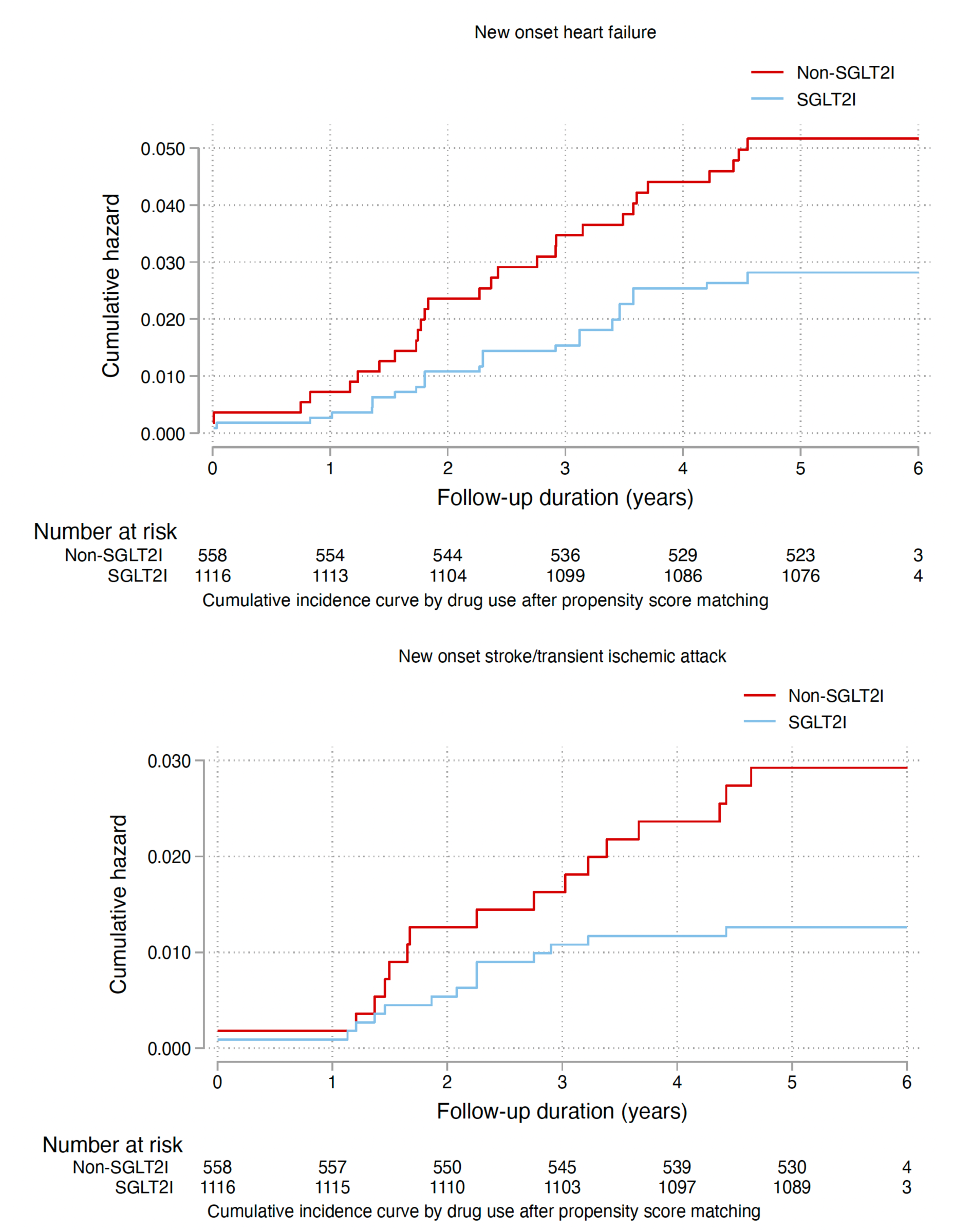
Cumulative incidence curves for new onset cardiovascular outcomes and all-cause mortality stratified by drug exposure effects of SGLT2I users and non-SGLT2I users after propensity score matching (1:2). SGLT2I: Sodium-glucose cotransporter-2 inhibitors;

**Table 2.** Incidence rate (IR) per 1000 person-year and multivariate Cox regression models of new onset cardiovascular disease and all-cause mortality in the cohort amongst GLP1A users before and after 1:2 propensity score matching. * for p≤ 0.05, ** for p ≤ 0.01, *** for p ≤ 0.001; CI: confidence interval; SGLT2I: sodium glucose cotransporter-2 inhibitor Adjusted for significant demographics, past comorbidities, duration of diabetes mellitus, and number of prior hospitalizations, number of anti-diabetic drug classes, other anti-diabetic medications, abbreviated MDRD, HbA1c, fasting glucose.

| **Myocardial infarction** | **Number of patients** | **Number of events** | **Total person-year** | **Incidence [95% CI] per 1000 person-time** | **Adjusted hazard ratio [95% CI]** | **P-value** |
| --- | --- | --- | --- | --- | --- | --- |
| **SGLT2I users** | 1116 | 21 | 6153.1 | 3.4[2.1-5.2] | 1 [Reference] | NA |
| **Non-SGLT2I users** | 558 | 22 | 3023.6 | 7.3[4.6-11.0] | 2.91[1.30-6.50] | 0.0092** |
| **Atrial fibrillation** | **Number of patients** | **Number of events** | **Total person-year** | **Incidence [95% CI] per 1000 person-time** | **Adjusted hazard ratio [95% CI]** | **P value** |
| **SGLT2I users** | 1116 | 13 | 6162.8 | 2.1[1.1-3.6] | 1 [Reference] | NA |
| **Non-SGLT2I users** | 558 | 13 | 3057.4 | 4.3[2.3-7.3] | 1.52[0.65-3.53] | 0.3335 |
| **Heart failure** | **Number of patients** | **Number of events** | **Total person-year** | **Incidence [95% CI] per 1000 person-time** | **Adjusted hazard ratio [95% CI]** | **P-value** |
| **SGLT2I users** | 1116 | 31 | 6131.3 | 5.1[3.4-7.2] | 1 [Reference] | NA |
| **Non-SGLT2I users** | 558 | 28 | 3014.8 | 9.3[6.2-13.4] | 2.49[1.22-5.08] | 0.0121* |
| **Stroke/ transient ischemic attack** | **Number of patients** | **Number of events** | **Total person-year** | **Incidence [95% CI] per 1000 person-time** | **Adjusted hazard ratio [95% CI]** | **P-value** |
| **SGLT2I users** | 1116 | 14 | 6165.8 | 2.3[1.2-3.8] | 1 [Reference] | NA |
| **Non-SGLT2I users** | 558 | 16 | 3048 | 5.2[3.05-8.5] | 1.72[0.70-4.24] | 0.2361 |
| **All-cause mortality** | **Number of patients** | **Number of events** | **Total person-year** | **Incidence [95% CI] per 1000 person-time** | **Adjusted hazard ratio [95% CI]** | **P value** |
| **SGLT2I users** | 1116 | 15 | 6215.3 | 2.4[1.4-4.0] | 1 [Reference] | NA |
| **Non-SGLT2I users** | 558 | 13 | 3097.8 | 4.2[2.2-7.2] | 1.66[0.61-4.52] | 0.3184 |

Besides, 13 SGLT2I users and 13 non-SGLT2I users developed atrial fibrillation. After a total follow-up of 9220.2 person-year, the incidence of atrial fibrillation was lower amongst SGLT2I users (IR per 1000 person-year: 2.1; 95% CI: 1.1-3.6) compared to non-SGLT2I users (IR per 1000 person-year: 4.3; 95% CI: 2.3-7.3) **(Table 2)**. Non-SGLT2I users did not had a significant risk of atrial fibrillation after adjustment (HR: 1.52; 95% CI: 0.65-3.53, p=0.3335) compared to SGLT2I users after adjustments **(Table 2)**. This was substantiated by the cumulative incidence curves (**Figure 2**).

Furthermore, 31 SGLT2I users and 28 non-SGLT2I users developed heart failure. After a total follow-up of 9146.1 person-year, the incidence of heart failure was lower amongst SGLT2I users (IR per 1000 person-year: 5.1; 95% CI: 3.4-7.2) compared to non-SGLT2I users (IR per 1000 person-year: 9.3; 95% CI: 6.2-13.4) **(Table 2)**. Non-SGLT2I users had a significant high risk of heart failure after adjustment (HR: 2.49; 95% CI: 1.22-5.08, p=0.0121) compared to SGLT2I users after adjustments **(Table 2).** This was substantiated by the cumulative incidence curves (**Figure 2**). The E-value for the heart failure was 4.42.

Finally, 14 SGLT2I users and 16 non-SGLT2I users developed stroke/TIA. After a total follow-up of 9213.8 person-year, the incidence of stroke/TIA was lower amongst SGLT2I users (IR per 1000 person-year: 2.3; 95% CI: 1.2-3.8) compared to non-SGLT2I users (IR per 1000 person-year: 5.2; 95% CI: 3.05-8.5) **(Table 2)**. Compared to SGLT2I users, non-SGLT2I users did not have significantly different risk of stroke/TIA after adjustment (HR: 1.72; 95% CI: 0.70-4.24, p=0.2361) **(Table 2)**. This was substantiated by the cumulative incidence curves (**Figure 2**).

### Association between SGLT2I and non-users and all-cause mortality

Overall, 15 SGLT2I users and 13 non-SGLT2I users passed away. After a follow-up of 9313.1 person-year, the incidence of all-cause mortality was lower amongst SGLT2I users (IR: 2.4; 95% CI: 1.4-4.0) compared to non-users (IR: 4.2; 95% CI: 2.2-7.2) **(Table 2)**. Non-SGLT2I users were not associated with higher risks of all-cause mortality after adjustment (HR: 1.66; 95% CI: 0.61-4.52, p=0.3184) compared to non-SGLT2I users after adjustments (**Table 2)**. This was substantiated by the cumulative incidence curves (**Figure 2**).

### Subgroup analysis

The results of the subgroup analysis for effects of SGLT2I and non-SGLT2I users on the cardiovascular outcomes are shown in **Figure 3**. The result demonstrated that non-SGLT2I users was associated with higher risks of myocardial infarction amongst patients without prior ischaemic heart disease. Besides, non-SGLT2I users were associated with higher risks of atrial fibrillation amongst female, with hypertension, hyperlipidaemia, on insulin, and on dipeptidyl peptidase-4 inhibitor (DPP4I). Furthermore, non-SGLT2I users were associated with higher risks of heart failure amongst female, with hypertension, hyperlipidaemia, without prior ischaemic heart disease, and on DPP4I. Lastly, non-SGLT2I users were associated with higher risks of stroke/TIA amongst patients with hypertension, on insulin, and without DPP4I.

**Figure 3.**
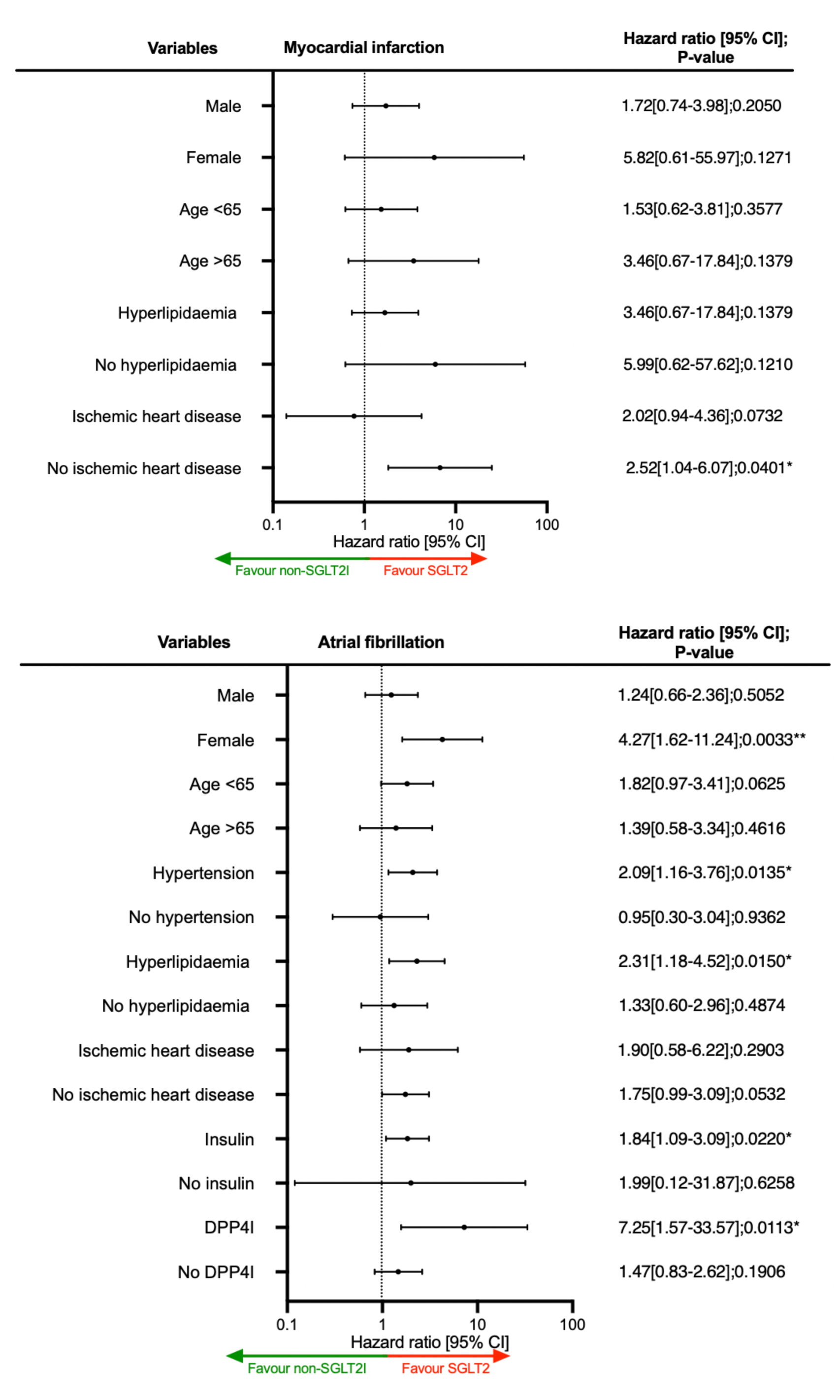
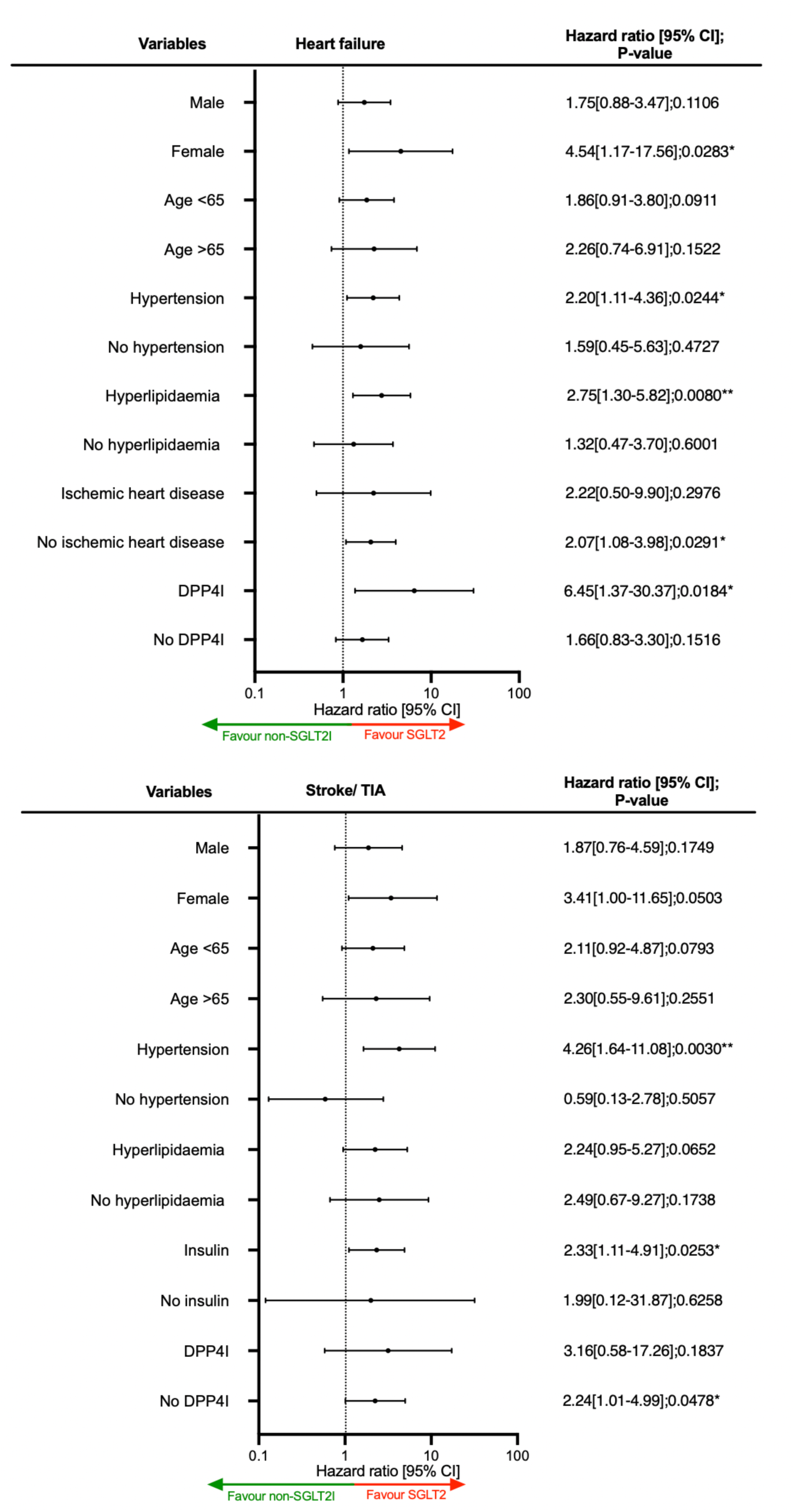
Forests plot of hazard ratios with 95% CI for SGLT2I users versus non-SGLT2I users on cardiovascular outcomes in the matched cohort. SGLT2I: Sodium-glucose cotransporter-2 inhibitors; DPP4I: Dipeptidyl peptidase-4 inhibitors; TIA: Transient ischaemic attack

The marginal effects analysis demonstrated that SGLT2I use was associated with lower risks of cardiovascular disease regardless of the time-weighted mean fasting glucose level apart from atrial fibrillation. Meanwhile, the differences between SGLT2I users and non-SLGT2I users on myocardial infarction, atrial fibrillation, and stroke/TIA became smaller as the time-weighted mean of HbA1c level increases (**Supplementary** Figure 2).

### Sensitivity analysis

Sensitivity analyses were performed to confirm the predictability of the models and test the potential effects on bias. The results of the cause-specific hazard models, sub-distribution hazard models and different propensity score approaches demonstrated that different models did not change the point estimates for both the myocardial infarction (all p<0.05) **(Supplementary Table 2)**. However, non-SGLT2I users were associated with a higher risk of stroke/TIA compared to SGLT2I users. The association between SGLT2I users and non-SGLT2I users on myocardial infarction and heart failure remained consistent after 1) excluding patients with less than 3-month follow-up duration; 2) Exclude patients with CKD stage 4/5 (eGFR <30), peritoneal dialysis or haemodialysis; 3) Excluding patients on financial aid; 4) Exclude patients at the top or bottom 5% of propensity score matching **(Supplementary Table 3)**.

### Machine learning causal inference analysis results

The causal inference analysis was performed using Casual Survival Forests model. The result demonstrated that SGLT2I demonstrated a higher estimated treatment effects (ETE) against myocardial infarction (Averaged treatment effect [ATE]: 0.009; 95% CI: 0.005-0.012; p=0.0012) and heart failure (ATE: 0.017; 95% CI: 0.012-0.023; p=0007), atrial fibrillation (ATE: 0.013; 95% CI: 0.009-0.018; p=0.0041), and stroke/TIA (ATE: 0.029; 95% CI: 0.022-0.032; p<0.0001) (**Figure 4**). Here ATE is calculated by ETE of SGLT2I minus ETE of non-SGLT2I. Furthermore, for the significant outcome myocardial infarction and heart failure in **Table 2**, in the causal inference analysis, SGLT2I users demonstrated a greater estimated treatment effects compared to non-SGLT2I users regardless of sex, hyperlipidaemia, hypertension, prior ischaemic heart disease and insulin **(Supplementary** Figure 3).

**Figure 4.**
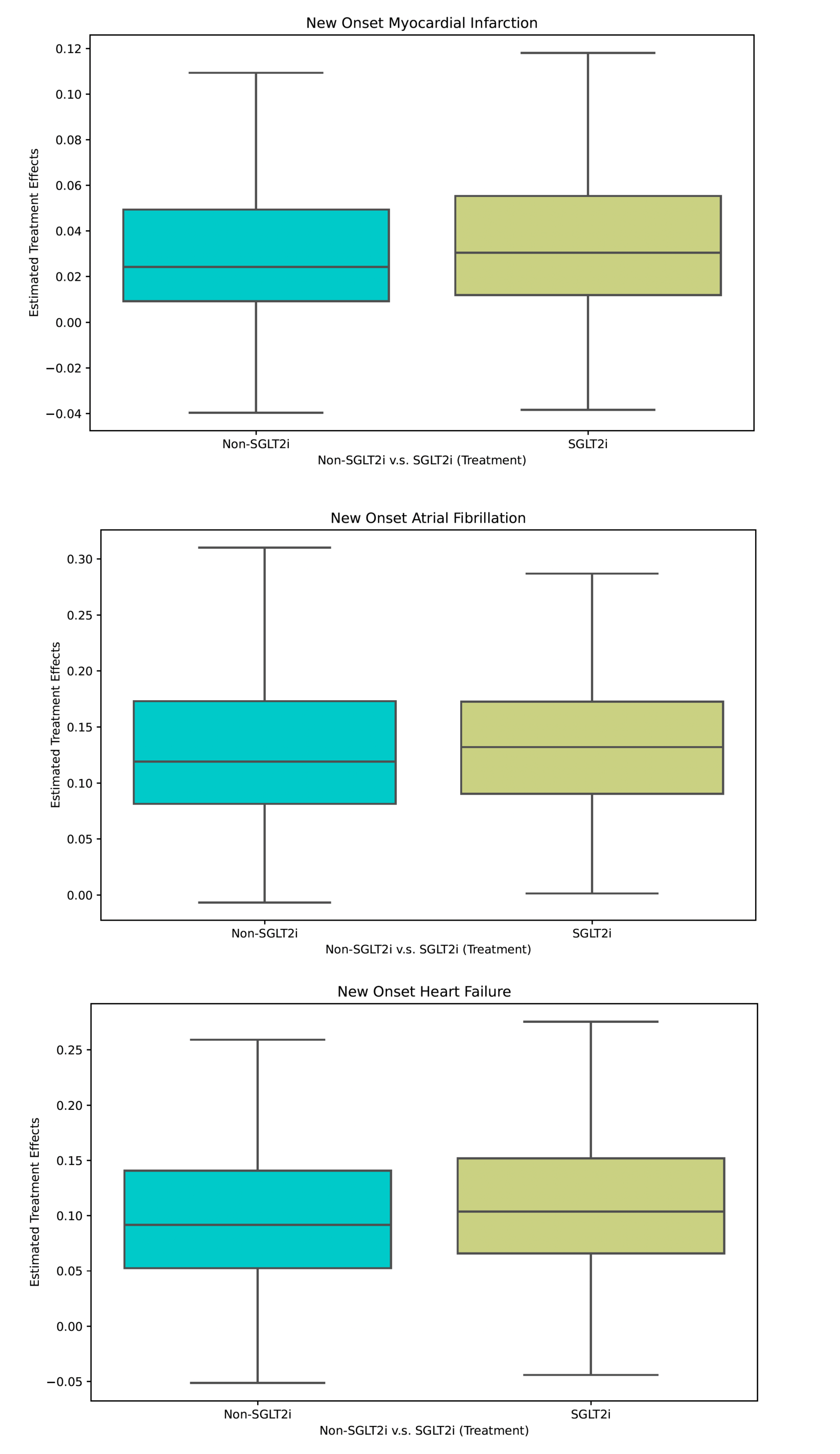
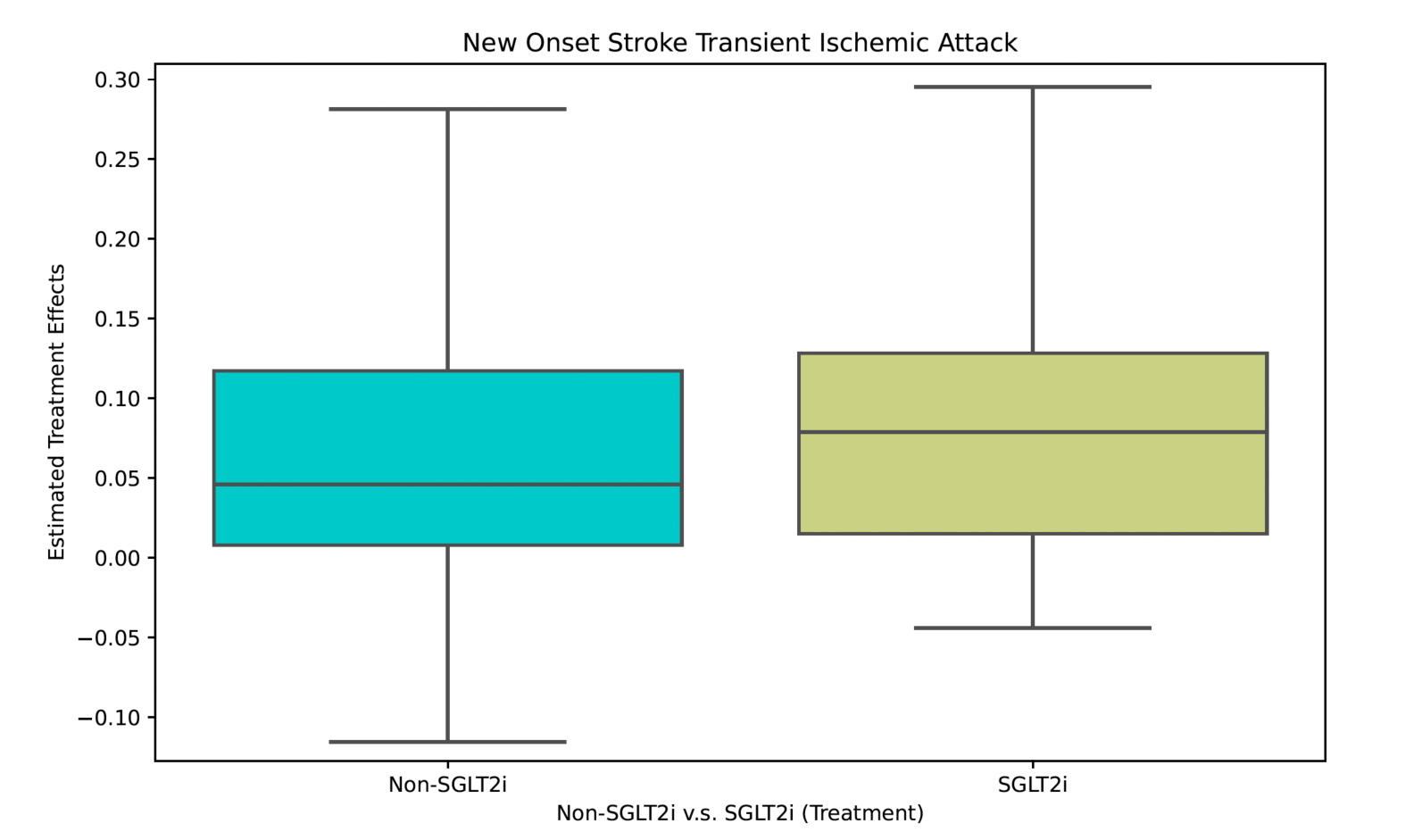
Treatment effect estimated by machine learning causal inference analysis. SGLT2I: Sodium-glucose cotransporter-2 inhibitors

### Falsification analysis

Hip fracture was used as negative control outcome in the falsification analysis for the comparison between SGLT2I and non-SGLT2I users **(Supplementary Table 4)**. The results demonstrated that non-SGLT2I users’ risk of the hip fracture after adjustment (HR: 0.91; 95% CI: 0.82-1.79, p=0.2311) were not significantly different from SGLT2I users.

## Discussion

In this territory-wide cohort study, real-world data was used to compare the association between SGLT2I usage and cardiovascular events amongst patients on GLP1a. The results suggest that non-SGLT2I users demonstrated a significantly higher risk of myocardial infarction and heart failure than SGLT2I users. Meanwhile, in the machine learning causal inference analysis, the result demonstrated that amongst patients on GLP1a, SGLT2I had a larger estimated treatment effect compared to non-SGLT2I users on cardiovascular outcomes.

### Comparison with previous studies

A recent consensus report jointly issued by the American Diabetes Association (ADA) and the European Association for the Study of Diabetes (EASD) has presented an extensive guideline endorsing the utilization of SGLT2I, as well as GLP-1A for individuals with T2DM with increased risk of cardiovascular disease and heart failure [22]. However, limited emphasis is placed on delineating the potential cardioprotective benefits arising from a combined therapy involving SGLT2I and GLP-1A.

In our study, we observed a notable increase in the risks of myocardial infarction and heart failure amongst SGLT2I non-users compared to the SGLT2I users in patients on GLP1a. The result remained consistent with a meta-analysis, which demonstrated that SGLT2I was associated with lower risks of heart failure compared to non-SGLT2I users, which was consistent with the clinical trial data [23]. Furthermore, in the meta-analysis including 97 trials, SGLT2I was associated with a lower risk of myocardial infarction compared to control with an HR of 0.86 [24]. SGLT2I has also demonstrated its protective effect against new-onset myocardial infarction compared to DPP4I in a Hong Kong population-based study (HR: 0.81) [25]. Yet, upon conversion, the equivalent HR for myocardial infarction amongst GLP1a patients in this study was 0.48, much lower than the published figures. This might indirectly imply that SGLT2I might exert greater protective effects amongst GLP1a patients. Nevertheless, evidence suggests SGLT2I was not significantly associated with a lower risk of acute coronary artery syndrome compared to non-SGLT2I users amongst elderly T2DM patients [26] and patients with renal failure [27]. These differences could be explained by the baseline atherosclerotic profile and the cardiorenal status; in this study, GLP1a alone, with or without interacting with SGLT2I, might have already improved those factors [28, 29].

While our findings corroborate the existing literature concerning the association between SGLT2I and a reduction in MACE, the underlying mechanistic pathways remain an imperative area for ongoing investigation. It was previously suggested that a combination of both SGLT2I and GLP1a could demonstrate superior glycaemic control with a greater reduction in the HbA1c compared to using either alone [30]. The associations between the use of SGLT2I and atrial fibrillation or stroke/TIA were not statistically significant. However, this could be explained by the relatively low case numbers for those outcomes.

The promise of the SGLT2I/GLP1a combination therapy in T2DM patients is evident in a post-hoc analysis derived from the EXSCEL trial; the result demonstrated that the combination of SGLT2I and GLP-1A yielded a decreased risk of MACE (HR: 0.68) compared to GLP-1A alone (HR: 0.85) [31]. Meanwhile, a US-based real-world cohort study showcased an overall risk reduction effect resulting from the addition of SGLT2I to the GLP1A regimen on MACE, but not for stroke [8]. This raised the possibility that SGLT2I might play a slightly lesser role in preventing stroke [32, 33].

The ML causal inference analysis was designed to distinguish between causal relationships and spurious correlations and would provide actionable information regarding the treatment [34]. The estimated treatment difference was defined as the difference in the predicted outcome for patients treated with that drug versus their outcome if they had not been treated [34]. This approach has been used to compare the effects of antidiabetic drugs amongst poorly controlled diabetes patients [35]. In our ML causal inference analysis, the result demonstrated that SGLT2I demonstrated higher estimated treatment effects against all 4 outcomes we investigated in this study. Besides, the causal ML method demonstrated that differences in outcomes with and without the usage of SGLT2I remained consistent across different subgroups, such that the effects of SGLT2I remained homogenous.

### Clinical implications

As the prevalence of T2DM continues to rise in Asia [36], the expanding body of literature concerning the potential cardioprotective benefits of antidiabetic drugs holds significant relevance. Under the current reference framework in Hong Kong designates metformin as the first-line therapy for T2DM [37, 38], with second-line antidiabetic drugs to be added when the glucose levels are insufficiently controlled. In our study, the number needed to treat for myocardial infarction and heart failure was 49 and 45, respectively **(Supplementary Table 5)**. While affirming the favourable cardioprotective effects of SGLT2I supplementation for GLP1a-treated T2DM patients, this study also underscores the necessity for further trials and in-depth investigations to ascertain whether these outcomes are solely attributable to SGLT2I or result from synergistic effect with GLP1a.

### Limitations

However, there are some limitations to the study. The observational nature of this study suggests that data on certain variables, namely alcohol use, BMI, smoking, family history and physical activity, could not be obtained and analysed. In compensation for this, laboratory results and comorbidities related to the cardiovascular outcomes were used to infer possible risk factors. To corroborate, drug cohorts were matched over a variety of medications and diseases, adjusted with regression, and performed sensitivity analyses to reduce the effect of the confounders.

The observational design of this study inherently lacks randomization, introducing potential biases. While propensity score matching was employed to mitigate confounding, the possibility of unidentified residual confounding remains. Therefore, a falsification analysis was conducted, and the result did not falsify the association observed. Additionally, the observational nature of the study precludes the establishment of definitive causal relationships. Yet, the incorporation of the machine learning approaches arguably holds promise in unravelling causal relationships, further accentuating the associations between SGLT2I usage and cardiovascular outcomes.

## Conclusion

This population-based cohort study suggested that non-SGLT2I users were associated with higher risks of myocardial infarction and heart failure compared to SGLT2I users amongst T2DM patients currently on GLP1a. These findings were substantiated by the machine learning causal inference analysis.

## Data Availability

Data are not available, as the data custodians (the Hospital Authority and the Department of Health of Hong Kong SAR) have not given permission for sharing due to patient confidentiality and privacy concerns. Local academic institutions, government departments, or nongovernmental organizations may apply for the access to data through the Hospital Authority's data sharing portal (https://www3.ha.org.hk/data).

## Funding source

The authors received no funding for the research, authorship, and/or publication of this article.

## Ethical approval statement

This study was approved by the Institutional Review Board of the University of Hong Kong/Hospital Authority Hong Kong West Cluster (HKU/HA HKWC IRB) (UW-20-250), and New Territories East Cluster-Chinese University of Hong Kong (NTEC-UCHK) Clinical Research Ethnics Committee (2018.309, 2018.643) and complied with the Declaration of Helsinki.

## Conflicts of Interest

None.

## Acknowledgements

None.

## Guarantor Statement

All authors approved the final version of the manuscript. GT is the guarantor of this work and, as such, had full access to all the data in the study and takes responsibility for the integrity of the data and the accuracy of the data analysis.

## Author contributions

Data analysis: ZYL, OHIC, JZ

Data review: ZYL, OHIC, CTC, GT, JZ

Data acquisition: OHIC, ZMWN, GT

Data interpretation: OHIC, CTC, JSKC, GT, JZ

Critical revision of manuscription: ZYL, RNCC, TTZ, TL, LL, JSKC, GT, JZ

Supervision: TTZ, TL, BMYC, GT, JZ

Manuscript writing: ZYL, OHIC, CTC, JSKC, JZ

Manuscript revision: ZYL, OHIC, CTC, JSKC, JZ

## Notes

### Competing Interest Statement

The authors have declared no competing interest.

