## Supplementary Appendix for "Compare SGLT2I versus non-SGLT2I users in type-2 diabetic mellitus patients on GLP-1 receptor agonist: A population-based and machine learning causal inference analysis"

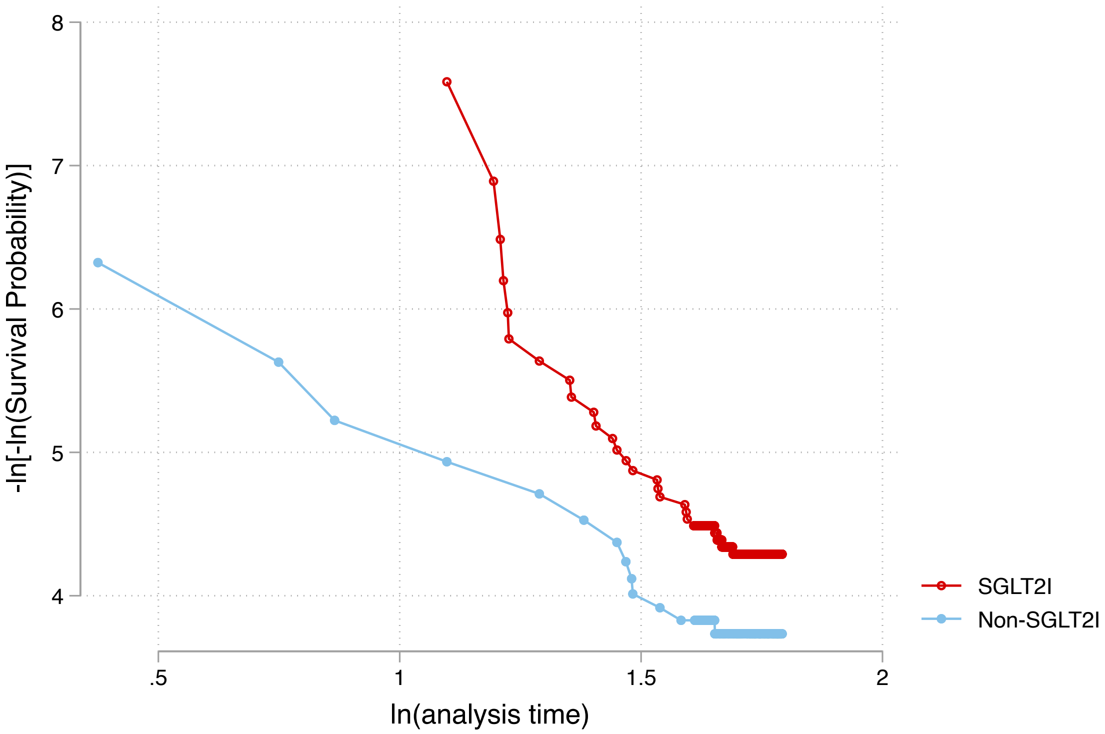

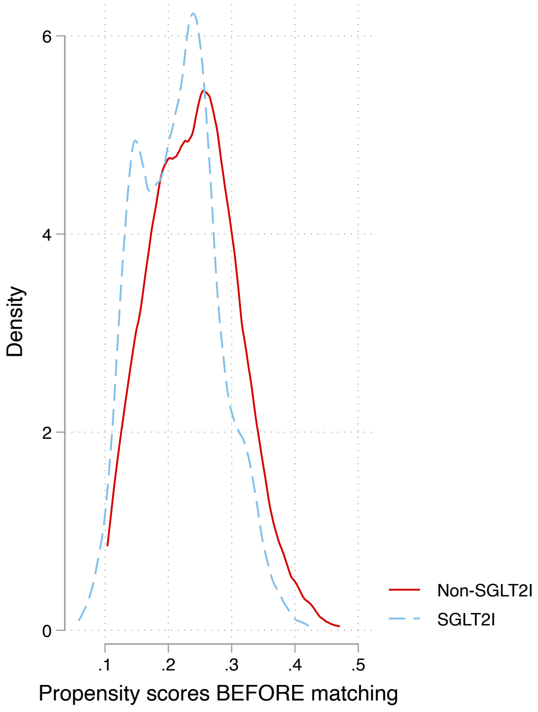

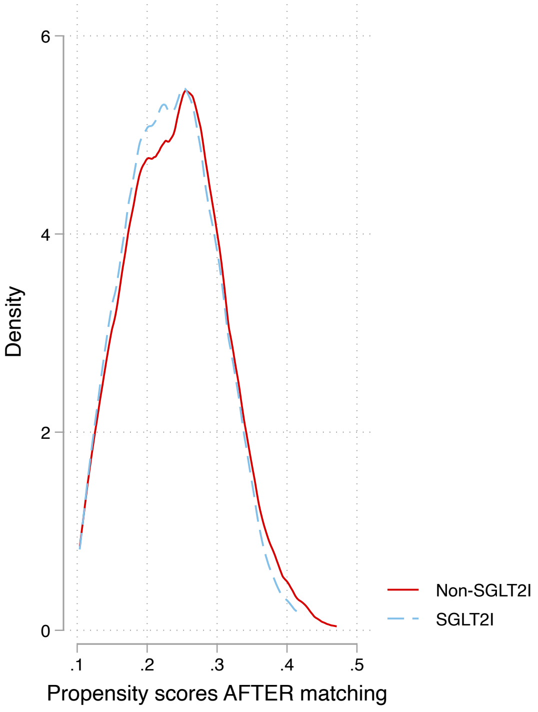

Nearest neighbor search strategy with caliper=0.1.

**Supplementary Figure 1. Propensity score matching comparisons and proportional hazard assumption checking with parallel lines for non-SGLT2I versus SGLT2I before and after 1:2 matching with nearest neighbour search strategy with calliper of 0.**

**Supplementary figure 2. Marginal effects of time-weighted mean fasting glucose and HbA1c with 95% CIs on cardiovascular outcomes in the matched cohort.**

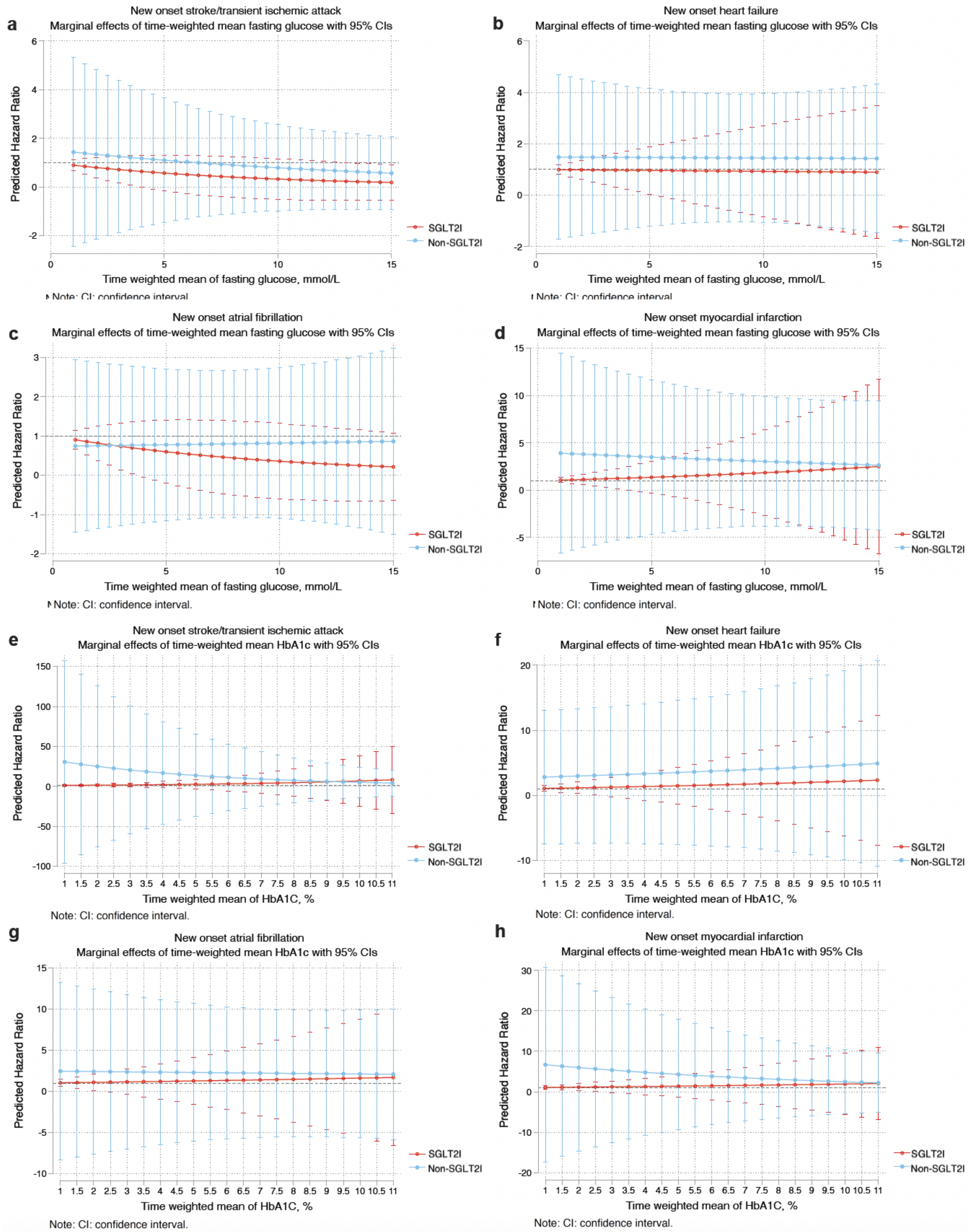

**Supplementary figure 3. Treatment effect estimated by machine learning causal inference analysis for myocardial infarction and heart failure across different subgroups.**

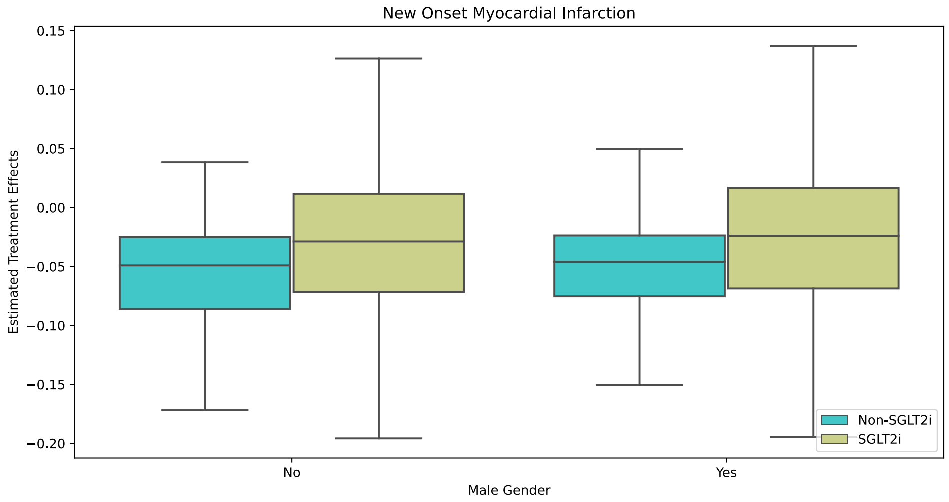

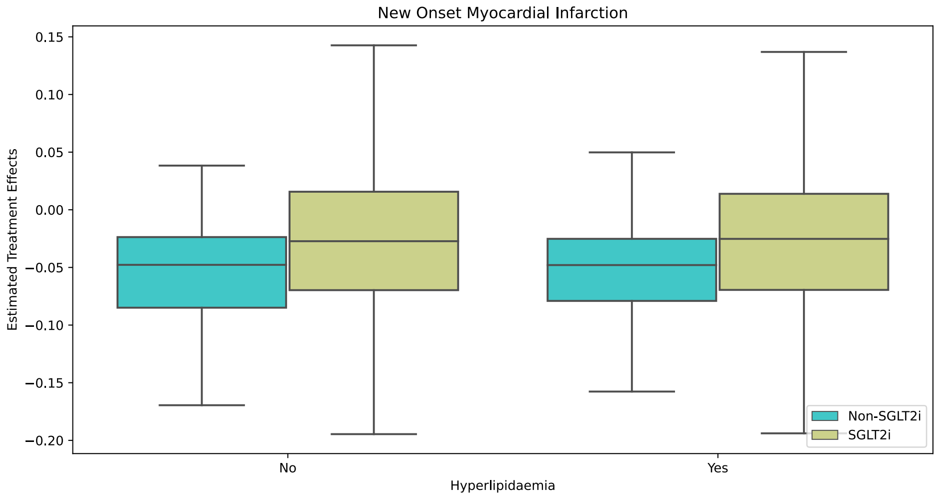

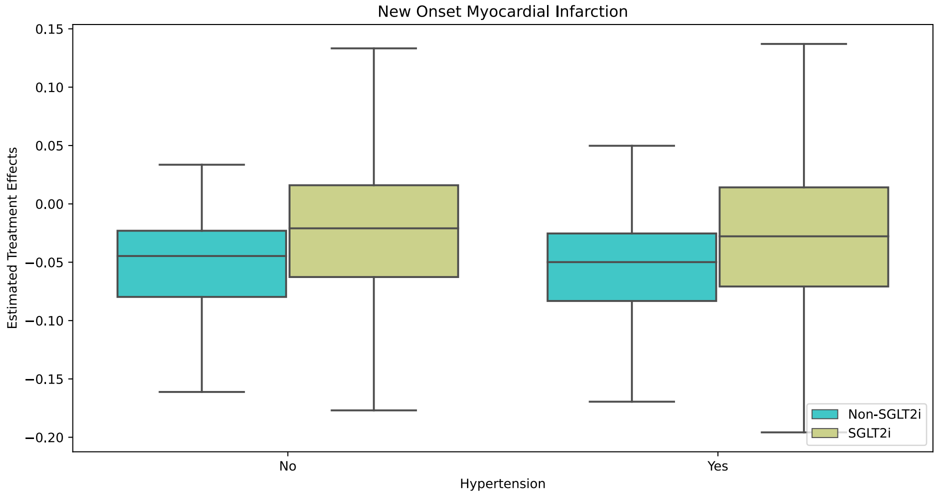

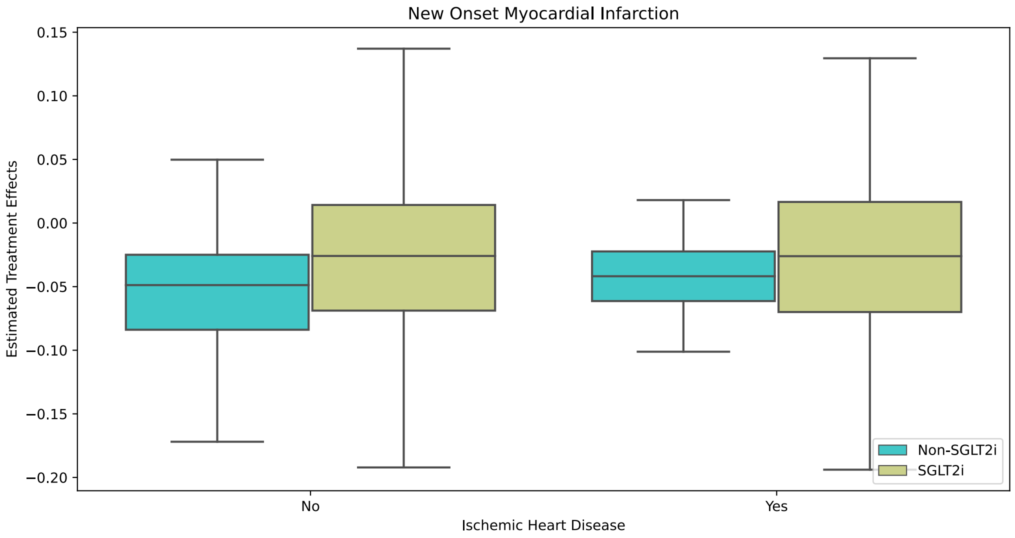
New Onset Myocardial Infarction
0.15
0.10
0.05
0.00
−0.05
−0.10
−0.15
−0.20
Estimated Treatment Effects
Non-SGLT2i
SGLT2i
No
Yes
Ischemic Heart Disease

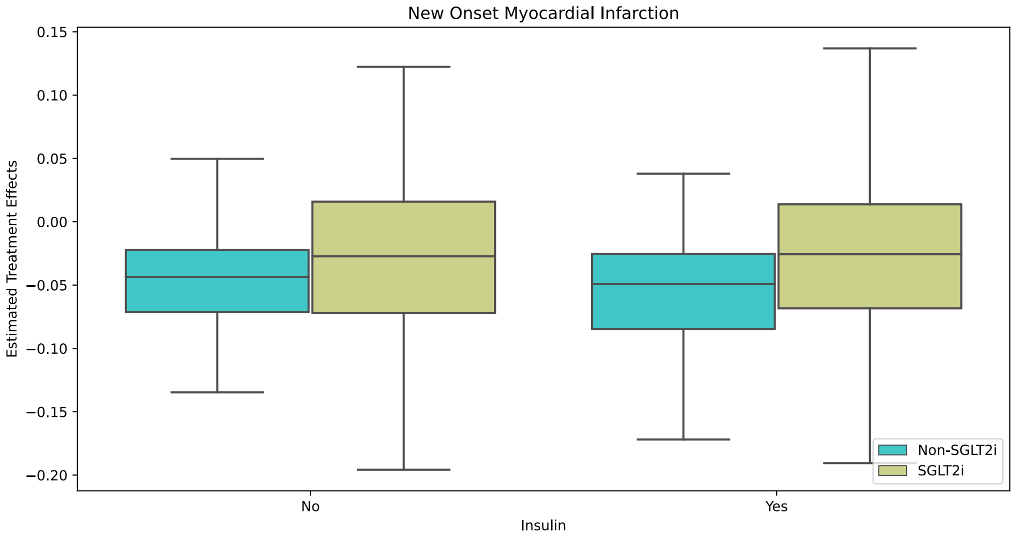
New Onset Myocardial Infarction
0.15
0.10
0.05
0.00
−0.05
−0.10
−0.15
−0.20
Estimated Treatment Effects
Non-SGLT2i
SGLT2i
No
Yes
Insulin

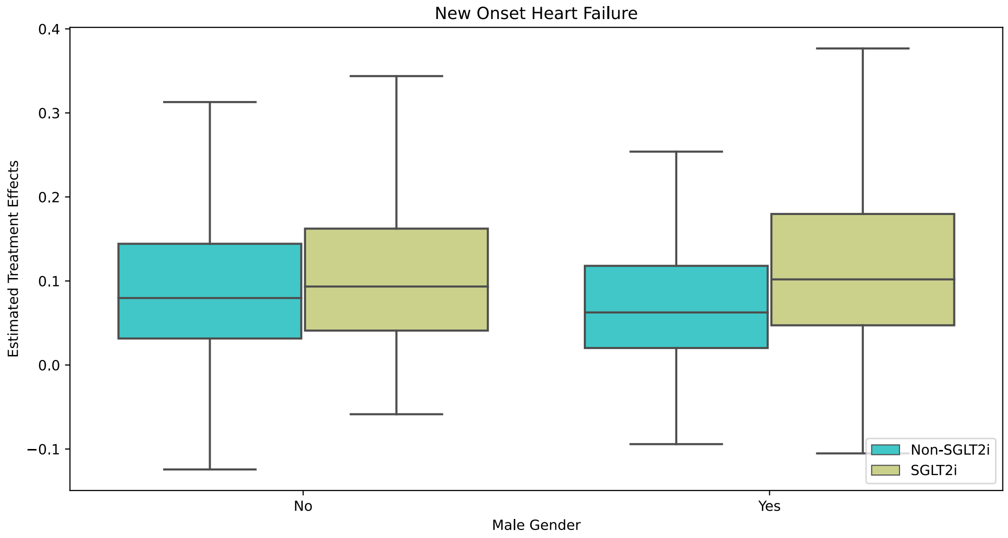
New Onset Heart Failure
0.4
0.3
0.2
0.1
0.0
−0.1
Estimated Treatment Effects
Non-SGLT2i
SGLT2i
No
Yes
Male Gender

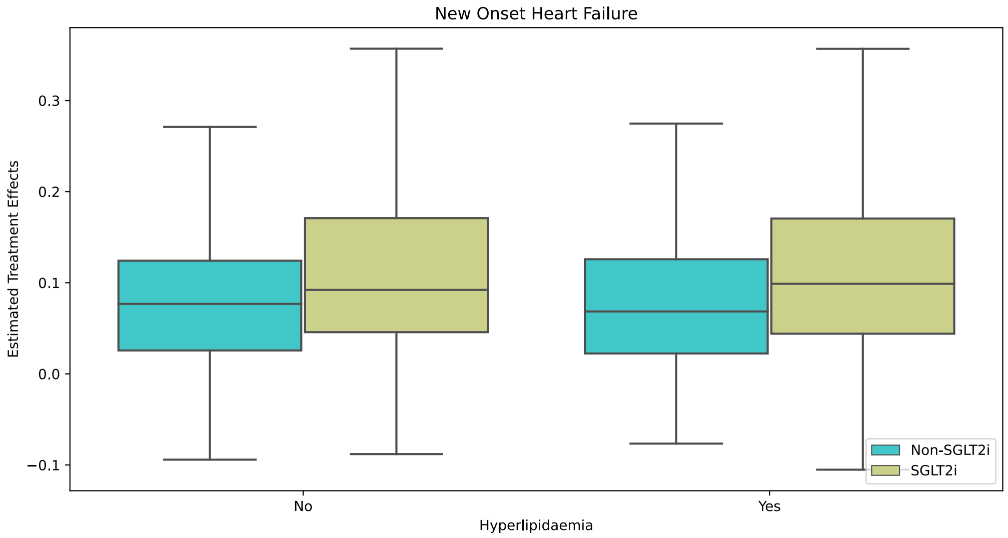
New Onset Heart Failure
Estimated Treatment Effects
0.3
0.2
0.1
0.0
−0.1
No
Yes
Hyperlipidaemia
Non-SGLT2i
SGLT2i

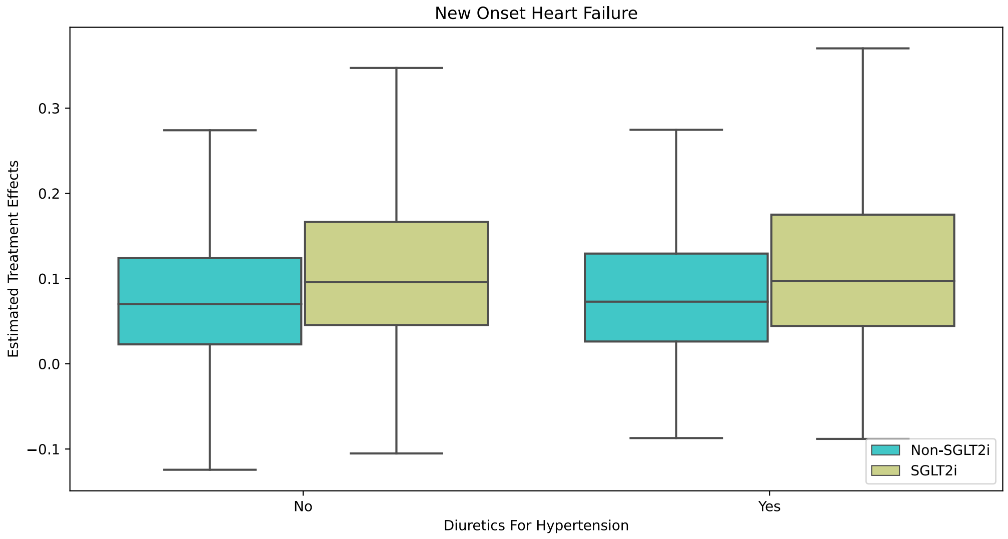
New Onset Heart Failure
Estimated Treatment Effects
0.3
0.2
0.1
0.0
−0.1
No
Yes
Diuretics For Hypertension
Non-SGLT2i
SGLT2i

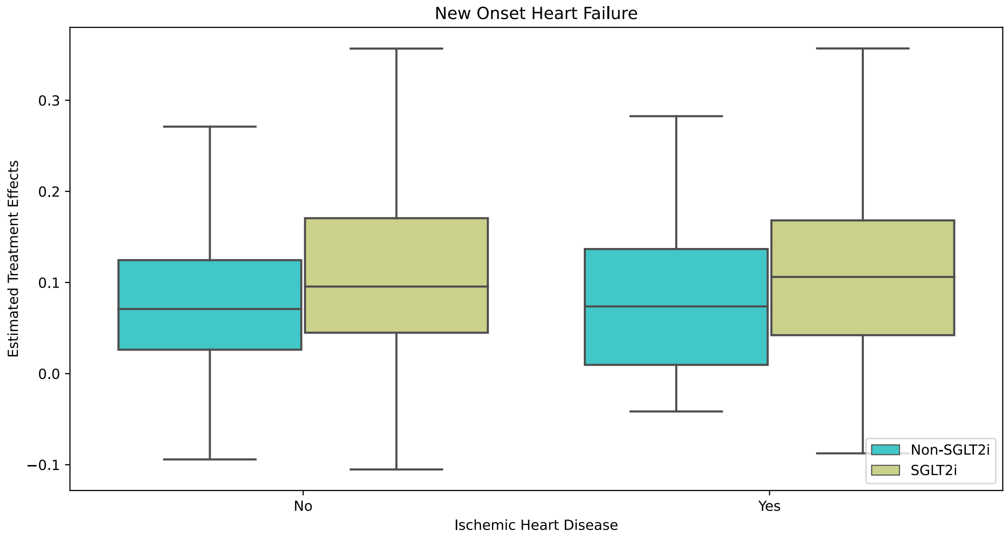
New Onset Heart Failure
Estimated Treatment Effects
0.3
0.2
0.1
0.0
−0.1
No
Yes
Ischemic Heart Disease
Non-SGLT2i
SGLT2i

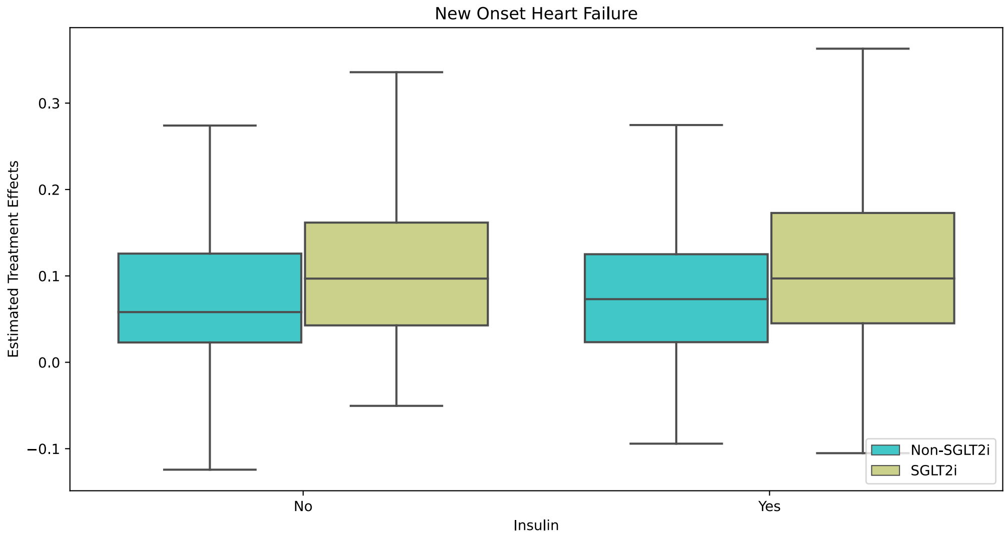
New Onset Heart Failure
Estimated Treatment Effects
0.3
0.2
0.1
0.0
−0.1
No
Yes
Insulin
Non-SGLT2i
SGLT2i

**Supplementary figure 4. The bias plot illustrating different combinations of RRUD and RREU for myocardial infarction and heart failure.**

RREU=strength of association between the unmeasured confounder and exposure; RRUD=strength of association between the unmeasured confounder and outcome

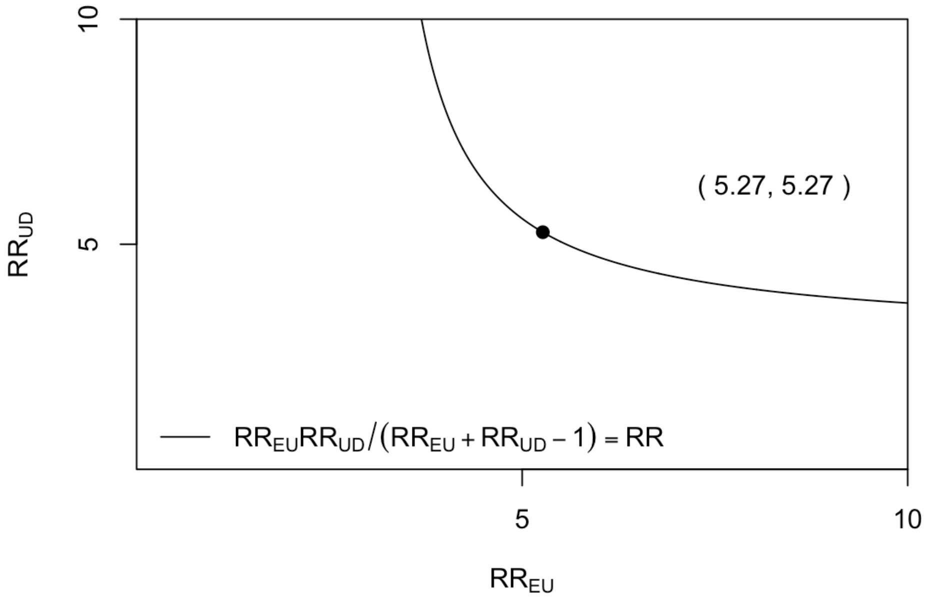

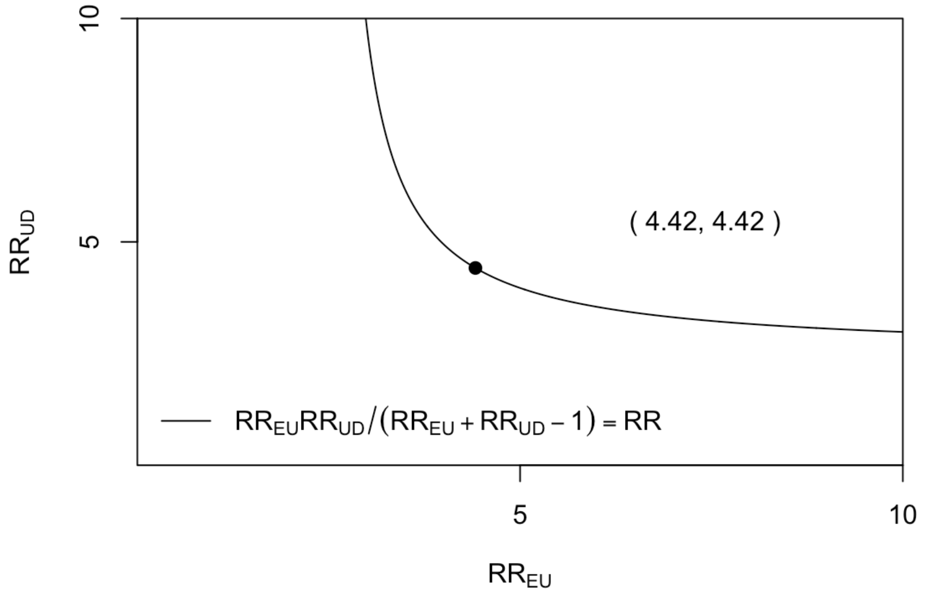

**Supplementary Table 1. The International Classification of Diseases, Clinical Modification (ICD-9-CM) codes for definitions of past comorbidities and outcomes.**

| Adverse outcome of interest |
|---|
| **Heart failure:** 428 428 428.1 428.2 428.2 428.21 428.22 428.23 428.3 428.3 428.31 428.32 428.33 428.4 428.4 428.41 428.42 428.43 428.9 398.91 402.01 402.11 402.91 404.01 404.03 404.11 404.13 404.91 404.93 |
| **Atrial fibrillation:** 427.31 429.4 |
| **Stroke/transient ischemic attack:** 435 435.1 435.2 435.3 435.8 435.9 433.81 433.91 434 436 437 437.1 433.31 433.01 434.01 434.1 434.11 434.9 434.91 437.2 437.3 437.4 437.5 437.6 437.7 437.8 437.9 430 431 432 432.1 432.9 |
| **Acute myocardial infarction:** 410 410.01 410.02 410.1 410.11 410.12 410.2 410.21 410.22 410.3 410.31 410.32 410.4 410.41 410.42 410.5 410.51 410.52 410.6 410.61 410.62 410.7 410.71 410.72 410.8 410.81 410.82 410.9 410.91 410.92 |
| **Past comorbidities** |
| **Cancer**: 140-239 |
| **Hypertension**: 401 401.1 401.9 402 402.01 402.1 402.11 402.9 402.91 403 403.01 403.1 403.11 403.9 403.91 404 404.01 404.02 404.03 404.1 404.11 404.12 404.13 404.9 404.91 404.92 404.93 405 405.01 405.09 405.1 405.11 405.19 405.9 405.91 405.99 437.2 + history of uses of anti-hypertensives |
| **Hyperlipidaemia**: 272.0 272.1 272.2 272.3 272.4 + history of uses of lipid-lowering drugs + lipid laboratory results |
| **Liver disease:** 275.1 275.0 572.0 572.4 572.1 572.3 572.8 573.0 573.4 573.8 573.9 |
| **Gastrointestinal bleeding:** 578.9 |
| **Ischemic heart disease:** 410.01 410.02 410.1 410.11 410.12 410.2 410.21 410.22 410.3 410.31 410.32 410.4 410.41 410.42 410.5 410.51 410.52 410.6 410.61 410.62 410.7 410.71 410.72 410.8 410.81 410.82 410.9 410.91 410.92 411 411.1 411.8 411.81 411.89 413 413.1 413.9 414 414.01 414.02 414.03 414.04 414.05 414.06 414.07 414.1 414.11 414.12 414.19 414.2 414.3 414.4 414.8 414.9 410 412 |
| **Renal diseases:** 582 582 582.1 582.2 582.4 582.8 582.81 582.89 582.9 583 583 583.1 583.2 583.4 583.6 583.7 585 585.1 585.2 585.3 585.4 585.5 585.6 585.9 586 588 588 588.1 588.8 588.81 588.89 588.9 |
| **Diabetic retinopathy:** 250.5 361 362.01 362.02 362.1 362.53 362.81 362.82 362.83 369 379.23 |
| **Diabetic nephropathy:** 250.40 250.41 250.42 250.43 |
| **Diabetic neuropathy**: 250.6, 337.0, 337.1, 354.0 – 355.9, 356.9, 357.2, 358.1, 536.3, 564.5, 596.54, 713.5, 951.0, 951.1, 951.3 |
| **Chronic obstructive pulmonary disease** 491.0 491.1 491.2 491.8 491.9 492.0 492.8 496 |
| **Venous thromboembolism:** 415.1 451, 453.1 453.2 453.3 453.8 453.9 |

**Supplementary Table 2. Sensitivity analyses for exposure effects of non-SGLT2I and SGLT2I on cardiovascular outcomes and all-cause mortality using different models.**

* for p≤ 0.05, ** for p ≤ 0.01, *** for p ≤ 0.001; SGLT2I: Sodium-glucose cotransporter-2 inhibitors; DPP4I: HR: hazard ratio; CI: confidence interval; PS: propensity score; IPTW: inverse probability of treatment weighting, SIPTW: stable inverse probability of treatment weighting.

| **Model** | **Myocardial infarction HR [95% CI];P value** | **Atrial fibrillation HR [95% CI];P value** | **Heart failure HR [95% CI];P value** | **Stroke/transient ischemic attack HR [95% CI];P value** | **All-cause mortality HR [95% CI];P value** |
|---|---|---|---|---|---|
| Cause-specific hazard models | 2.34[1.06-3.66];0.0311* | 1.98[0.88-3.51];0.1523 | 1.77[0.95-2.45];0.0623 | 2.43[1.15-4.99];0.0245* | 1.41[0.77-3.61];0.4511 |
| Sub-distribution hazard models | 2.65[1.12-3.98];0.0461* | 1.68[0.72-4.12];0.1522 | 1.98[0.78-2.67];0.0712 | 1.98[1.45-4.99];0.0245* | 1.66[0.89-3.98];0.3522 |
| PS stratification | 1.98[1.23-5.23];0.0254* | 1.98[0.88-3.51];0.1523 | 1.77[0.95-2.45];0.0623 | 2.43[1.15-4.99];0.0245* | 1.41[0.77-3.61];0.4511 |
| PS with IPTW | 2.77[1.09-4.55];0.0412* | 2.01[0.78-4.14];0.2364 | 1.89[0.93-2.88];0.0412* | 2.67[1.13-4.12];0.0109* | 1.56[0.56-2.89];0.5611 |
| PS with SIPTW | 3.12[1.24-4.89];0.0461* | 2.45[0.83-3.99];0.3551 | 1.38[0.88-2.65];0.0671 | 2.67[1.52-4.56];0.0156* | 1.56[0.82-3.88];0.4021 |

**Supplementary Table 3. Sensitivity analysis of SGLT2I users versus non-SGLT2I users on adverse outcomes.**

* for p≤ 0.05, ** for p ≤ 0.01, *** for p ≤ 0.001; SGLT2I: Sodium-glucose cotransporter-2 inhibitors; HR: hazard ratio; CI: confidence interval.

| **Model** | **All-cause mortality HR [95% CI];P value** | **Myocardial infarction HR [95% CI];P value** | **Atrial fibrillation HR [95% CI];P value** | **Heart failure H R [95% CI];P value** | **Stroke/transient ischemic attack HR [95% CI];P value** |
|---|---|---|---|---|---|
| Exclude patients with less than 3-month follow-up duration | 1.74[0.83-3.66];0.1428 | 2.13[1.17-3.87];0.0133* | 2.01[0.93-4.34];0.0743 | 1.83[1.10-3.06];0.0199* | 2.31[1.13-4.73];0.0223* |
| Exclude patients with MDRD <30 | 1.69[0.81-3.56];0.1646 | 2.19[1.16-4.14];0.0158* | 1.95[0.91-4.21];0.0879 | 2.24[1.26-3.98];0.0057** | 1.81[0.85-3.86];0.1219 |
| Exclude patients on Financial aid | 1.74[0.83-3.66];0.1427 | 2.24[1.22-4.10];0.0092** | 2.18[1.00-4.78];0.0513 | 1.76[1.04-2.97];0.0353* | 2.49[1.20-5.17];0.0147* |
| Exclude patients at the top or bottom 5% of propensity score matching | 1.74[0.83-3.66];0.1428 | 2.13[1.17-3.87];0.0133* | 2.01[0.93-4.34];0.0743 | 1.83[1.10-3.06];0.0199* | 2.31[1.13-4.73];0.0223* |

**Supplementary Table 4. Falsification analysis: Exposure effects of non-SGLT2I v.s. SGLT2I on new onset hip fracture in the matched cohort with 1:2 ratio.**

* for p≤ 0.05, ** for p ≤ 0.01, *** for p ≤ 0.001; SGLT2I: Sodium-glucose cotransporter-2 inhibitors; HR: hazard ratio; CI: confidence interval.

| | **Number of events, Count(%)** | **Time to new onset hip fractures, days (IQR)** | **New onset hip fractures<br>HR[95% CI];P value** |
|---|---|---|---|
| Non- SGLT2I (N=558) | 6(1.08%) | 2019(1945-2131) | 0.91[0.82-1.79];0.2311 |
| SGLT2I (N=1116) | 13(1.17%) | 2028(1934-2142) | 1.0[Reference] |

**Supplementary Table 5. Absolute Risk Reduction and Number Needed to Treat for of SGLT2I users versus non-SGLT2I users on adverse outcomes.**

SGLT2I: Sodium-glucose cotransporter-2 inhibitors.

| **Characteristics** | **Non-SGLT2i (Treatment control) (N=558)** | **SGLT2i (N=1116)** | **Absolute Risk Reduction (ARR)** | **Number Needed to Treat (NNT)** |
|---|---|---|---|---|
| All-cause mortality | 2.32% | 1.34% | 0.98% | 102 |
| Cardiovascular mortality | 0.53% | 0.26% | 0.27% | 370 |
| New onset myocardial infarction | 3.94% | 1.88% | 2.06% | 49 |
| New onset atrial fibrillation | 2.32% | 1.16% | 1.16% | 86 |
| New onset heart failure | 5.01% | 2.77% | 2.24% | 45 |
| New onset stroke/transient ischemic attack | 2.86% | 1.25% | 1.61% | 62 |
